# Isolated prolapse of the posterior mitral valve leaflet: phenotypic refinement, heritability and genetic etiology

**DOI:** 10.1101/2024.10.16.24315096

**Authors:** Antoine Rimbert, Damien Duval, Daniel Trujillano, Florence Kyndt, Antoine Jobbe-Duval, Pierre Lindenbaum, Nathan Tucker, Simon Lecointe, Pauline Labbé, Claire Toquet, Matilde Karakachoff, Jean-Christian Roussel, Christophe Baufreton, Patrick Bruneval, Caroline Cueff, Erwan Donal, Richard Redon, Robert Olaso, Anne Boland, Jean-François Deleuze, FranceGenRef Consortium, Xavier Estivill, Susan Slaugenhaupt, Roger R Markwald, Russel A Norris, Jean-Philippe Verhoye, Vincent Probst, Albert Hagège, Robert Levine, Xavier Jeunemaitre, Hervé Le Marec, Romain Capoulade, Nabila Bouatia-Naji, Christian Dina, David Milan, Stephan Ossowski, Jean-Jacques Schott, Jean Mérot, Solena Le Scouarnec, Thierry Le Tourneau

## Abstract

**BACKGROUND:** Isolated posterior leaflet mitral valve prolapse (PostMVP), a common form of MVP, often referred as fibroelastic deficiency, is considered a degenerative disease. PostMVP patients are usually asymptomatic and often undiagnosed until chordal rupture. The present study aims to characterize familial PostMVP phenotype and familial recurrence, its genetic background, and the pathophysiological processes involved.

**METHODS:** We prospectively enrolled 284 unrelated MVP probands, of whom 178 (63%) had bi-leaflet MVP and 106 had PostMVP (37%). Familial screening within PostMVP patients allowed the identification of 20 families with inherited forms of PostMVP for whom whole genome sequencing was carried out in probands. Functional *in vivo* and *in vitro* investigations were performed in zebrafishand in Hek293T cells.

**RESULTS:** In the 20 families with inherited form of PostMVP, 38.8% of relatives had a MVP/prodromal form, mainly of the posterior leaflet, with transmission consistent with an autosomal dominant mode of inheritance. Compared with control relatives, PostMVP family patients have clear posterior leaflet dystrophy on echocardiography. Patients with PostMVP present a burden of rare genetic variants in *ARHGAP24. ARHGAP24* encodes the filamin A binding RhoGTPase-activating protein FilGAP and its silencing in zebrafish leads to atrioventricular regurgitation. *In vitro* functional studies showed that variants of FilGAP, found in PostMVP families, are *loss-of-function* variants impairing cellular adhesion and mechano-transduction capacities.

**CONCLUSIONS:** PostMVP should not only be considered an isolated degenerative pathology but as a specific heritable phenotypic trait with genetic and functional pathophysiological origins. The identification of *loss-of-function* variants in *ARHGAP24* further reinforces the pivotal role of mechano-transduction pathways in the pathogenesis of MVP.

**CLINICAL PERSPECTIVE:**

- Isolated posterior mitral valve prolapse (PostMVP), often called fibro-elastic deficiency MVP, is at least in some patients, a specific inherited phenotypic trait
- PostMVP has both genetic and functional pathophysiological origins
- Genetic variants in the *ARHGAP24* gene, which encodes for the FilGAP protein, cause progressive Post MVP in familial cases, and impair cell adhesion and mechano-transduction capacities

## INTRODUCTION

Mitral valve prolapse (MVP), defined by an excessive leaflet displacement towards the left atrium in systole^1^, results in an inappropriate apposition of valve leaflets which contributes to mitral regurgitation (MR) and can lead to heart failure^2^. MVP affects 2 to 3% of the general population and is one of the most frequent indications for cardiac valve surgery in Western countries^1–3^. MVP is a heterogeneous group of disorders with a broad spectrum of clinical, echocardiographic, surgical and histological presentations^4–6^.

The most common form of MVP is Barlow disease (referred as bi-leaflet prolapse [BiMVP] in the present study), and is characterized by an early onset of the disease, a bi-leaflet elongation and prolapse with diffuse excess of leaflet tissue and an enlarged mitral annulus^5^. In contrast, isolated posterior mitral valve leaflet prolapse (PostMVP), often referred as fibroelastic deficiency (FED), displays a later onset, with localized valvular dystrophy generally restricted to one segment and moderate mitral annulus enlargement. In PostMVP, severe MR results from chordal rupture, located mainly on the median part of the posterior leaflet. In contrast with BiMVP patients, PostMVP patients are usually asymptomatic and remain undetected until they develop severe MR^3,5,7,8^. In the past decades the diagnosis criteria of typical and prodromal forms of MVP have greatly improved. Prodromal forms have been evidenced in some MVP families and strengthen the idea that MVP encompasses a more complex phenotypic expression than previously thought^3,5,7,8^. Although these prodromal forms have no, or little, immediate clinical impact, their diagnosis has substantial importance for genetic investigations. In addition, it is worth noting that these prodromal forms may progress to more severe forms during life^7^, and could be monitored periodically.

MVP has long been considered a degenerative disease until the discovery of inherited forms of MVP^9–11^ and the identification of genetic bases (*FLNA, DCHS1, DZIP1* as well as risk *loci* through genome wide association studies^12–17^). The combination of genetic evidence with deep-phenotyping of patients, together with the development of cellular and animal models, led to the understanding of some pathophysiological and molecular mechanisms involved in the pathogenesis of MVP in human (*e.g*. cellular mechanical stress response, extracellular matrix remodeling, primary cilia-dependent regulation and immune cell response^13,15,18–23^).

While molecular bases have been identified in non-syndromic forms of BiMVP, biological mechanisms involved and/or the exclusive degenerative nature of PostMVP remain elusive and debated.

In the present work, from 284 unrelated MVP patients (178 BiMVP and 106 PostMVP) prospectively enrolled in a genetic study, we offered family screening to relatives of PostMVP patients to 1) determine the sporadic or familial expression of PostMVP and its clinical presentation, 2) characterize its inheritance mode and genetic background and, 3) understand involved biological and pathophysiological processes.

## METHODS

### Patients

We prospectively enrolled 284 unrelated patients with non-syndromic MVP who underwent a comprehensive clinical and echocardiographic examination. Patients were classified as BiMVP patients and PostMVP patients. Familial screening was proposed to relatives of PostMVP cases. Blood was sampled for genetic studies in probands but also in all adult-relatives who accepted to participate in familial investigations. In children, salivary sampling was proposed for genetic studies. The study complies with the declaration of Helsinki, the French guidelines for genetic research and was approved by local ethic committees (Genetic and Phenotypic Characteristics of Mitral Valve Prolapse; NCT03884426). We obtained a favorable opinion from the local ethics body GNEDS (Groupe Nantais d’Ethique dans le Domaine de la Santé) on December 24, 2012 and a written informed consent was obtained from each participant.

### Echocardiography

Echocardiograms were carried out by experienced investigators using a commercially available digital ultrasound system (Vivid, GE Medical Systems, Milwaukee, WI, USA) and analyzed with a dedicated software (EchoPac PC; GE Healthcare, Waukesha, Wisconsin). Standard echocardiographic parameters were measured according to current recommendations^24^ as detailed in **Supplementary appendix**. The mitral valve (MV) apparatus was comprehensively assessed on echocardiography as previously described^8,25^. MVP was defined as a displacement of at least one leaflet ≥ 2 mm above the mitral annulus line, and a prodromal form as the presence of MV billowing < 2 mm^8^. Mitral regurgitation was quantified by 2 methods (proximal isovelocity surface area, volumetric method) and classified as trace/mild or moderate/severe according to a regurgitant volume < or ≥ 45 mL/beat, respectively.

### Genetic analyses

Genomic DNA was extracted from peripheral blood lymphocytes by standard protocols and whole genome sequencing (WGS) was performed at the Centre National de Recherche en Génomique Humaine (CNRGH, Institut de Biologie François Jacob, CEA, Evry, France). The experimental and analysis workflows are described in the **Supplementary appendix**. Briefly, we focused our analysis on rare variants affecting protein sequence (in coding or splicing genomic regions) which pass quality controls for 20 PostMVP probands of PostMVP families and 856 controls from a general healthy French population (FranceGenRef consortium). We tested the enrichment of selected variants in patients when compared to controls using the cohort allelic sums test (CAST)^26^. CAST test was applied by gene and raw not-adjusted p-values were computed. The validation of identified variants and their co-segregation with the PostMVP phenotype in families were performed by Sanger sequencing.

### *In vivo* experiments

All experiments were performed in accordance with approved Institutional Animal Care and Use Committee (IACUC) protocols at the Cardiovascular Research Center at MGH in Boston. Zebrafish of the Tubingen/AB strain were reared according to standard techniques. Minimal effective doses of antisense morpholino oligonucleotides were calculated by serial dilution of morpholino and evaluation of transcript expression at 96 hours post fertilization. Semi-quantitative PCR was used to demonstrate morpholino knockdown efficacy with ef-1a used as a reference gene. Effective doses were injected into single-cell stage embryos, manually dechorionated 24 hours post fertilization and treated with 0.003% phenylthiourea to inhibit pigmentation. 96 hours post fertilization, embryos were scored for presence of atrioventricular valve-regurgitation using high-speed videography (125fps) on a Nikon Eclipse 50i microscope coupled to a Fastec Imaging Inline IN250 high speed CCD camera.

### *In vitro* experiments

*In vitro* experiments were performed using Hek293T cells, culture, transfection conditions as well as plasmids and siRNA used in the present study, are detailed in **Supplementary appendix**.

#### Monitoring of cellular adhesion and spreading phenotypes

The monitoring of cell adhesion and spreading has been assessed with the xCELLigence system as previously described^19,27^ and detailed in **Supplementary appendix**. Briefly, the xCELLigence technology is a cellular biosensor, which measures the adhesion and spreading of cells to high-density gold electrode arrays printed on custom-designed plates. Variations in electrical impedance monitor cellular adhesion and spreading changes in real-time.

#### Evaluation of the GTPase activating properties (GAP) of FilGAP

FilGAP, encoded by the gene *ARHGAP24*, is a natural inhibitor of the Rac1-GTPase activity, involved, among others, in actin cytoskeleton remodeling. Its GTPase activating properties (GAP) activity catalyzes the hydrolysis of Rac1-GTP (Rac1 activated form (Rac1-GTP) into its inactivated form (Rac1-GDP). To evaluate the impact of the mutations on the GAP capacities of FilGAP we ran a Rac1-GAP activity assay (detailed in **Supplementary appendix).** Briefly, the activated form of Rac1 from cells transfected with WT or mutated forms of FilGAP was isolated in pull down assays using Glutathione-S-transferase (GST) and quantified by western blotting. The ratios Rac1-GTP / Rac1-total was used as an instrument to assess the effects of the mutations FilGAP activity^28^.

#### Co-immunoprecipitation and immunoblotting

Cells were co-transfected with HA and EGFP-tagged forms of FilGAP (mutated or wild-type (WT)) and immunoprecipitation of FilGAP-HA was performed using anti-HA-antibodies coated beads (Dynabeads, Invitrogen). Western blot analysis was performed to reveal co-immunoprecipitated proteins (detailed in **Supplementary appendix**).

### Statistical analysis

Baseline results are expressed as mean (±standard deviation) or numbers (percentage), as appropriate. Comparisons between groups were performed with Student’s t-test for continuous normally distributed parameters or a Wilcoxon test otherwise. Categorical parameters were compared with a chi-squared test or a Fisher’s exact test, when appropriate. The prevalence of atrioventricular regurgitation in zebrafish was compared using Fisher’s exact test. Results from *in vitro* experiments were analyzed using a Mann-Whitney test or two-ways ANOVA test: *, p < 0.05; **, p < 0.01; ***, p < 0.001. Two-tailed p-values < 0.05 were considered statistically significant. All analyses were performed with SPSS software version 19 (SPSS, Chicago, IL, USA), R version 3.3.2, or Prism (GraphPad Software). Specific statistical tests were applied in the genetic part and described in **Supplementary appendix.**

## RESULTS

### Patients

Baseline characteristics of the 284 unrelated patients with non-syndromic MVP are shown in **Table 1**. Mean age of the 284 patients was 58±14 years, 189 patients (67%) were males, 178 (63%) patients had BiMVP and 106 (37%) PostMVP. Mitral regurgitation was moderate/severe in most patients (85%). Among these 284 patients, clinical examination found only benign extra-cardiac abnormalities. Myopia was observed in 39 (14%) patients, narrow palate in 21 (7%), moderate scoliosis in 7 (2%), benign joint hypermobility in 13 (5%), pectus excavatum in 2 (1%) and flat feet in 13 (5%) patients.

**Table 1:** Baseline characteristics of the MVP cases. *Abbreviations:* MVP: mitral valve prolapse, AF: atrial fibrillation, SBP: Systolic blood pressure, BMI: body mass index, BiMVP: Bi-leaflet MVP, PostMVP: isolated posterior leaflet prolapse, MR: mitral regurgitation, ERO: effective regurgitant orifice area.

|  | MVP cases (n=284) |
| --- | --- |
| Age, years | 58±14 |
| Male, n (%) | 189 (67) |
| Heart rate, beat/min | 70±13 |
| AF, n (%) | 24 (8) |
| SBP, mmHg | 132±17 |
| Obesity (BMI ≥ 30), n(%) | 17 (6) |
| Hypertension, n (%) | 106 (37) |
| Diabetes, n (%) | 16 (6) |
| Dyslipidemia, n (%) | 77 (27) |
| Tobacco, n (%) | 86 (30) |
| BiMVP, n(%) | 178 (63) |
| PostMVP, n(%) | 106 (37) |
| MR grade, n(%) |  |
| -trace/mild | 42 (15) |
| -moderate/severe | 242 (85) |
| ERO, mm<sup>2</sup> | 37±21 |
| Regurgitant volume, mL | 57±32 |

### Familial screening and identification of inherited forms of PostMVP

From 106 patients with PostMVP, 30 accepted a familial screening. Within the families screened, we identified 20 PostMVP families with at least one PostMVP-affected relative (with 2 to 6 Post-MVP patients per family) (**Fig. S1 and S2**). In these families, the mode of inheritance of PostMVP was compatible with autosomal dominant transmission. Moreover, we identified a pair of monozygous twins, both presenting with a PostMVP (**Fig. S2**, Fam.9 II:3 and II:4) which strengthens the hypothesis of a genetic origin of PostMVP.

Overall, 136 individuals from the 20 PostMVP families were examined, of whom 65 (48%; inclusing 20 probants and 45 relatives) had at least a prodromal form of PostMVP (**Fig. S2**). In addition to the 20 probands with typical PostMVP, 24 (20.7%) relatives had MVP and 21 (18.1%) a prodromal MVP with posterior leaflet billowing giving an overall recurrence rate in relatives of 38.8%. Baseline clinical and echocardiographic characteristics of the 136 individuals are shown in **Table 2**. Patients with MVP or prodromal forms (n=65) were compared with control relatives (n=71). For MV apparatus phenotype characterization, only individuals > 16 years old (n=120) were considered for quantification. Patients with MVP were older (P=0.0002) and more susceptible to have hypertension (P=0.01) and dyslipidemia (P=0.03) than control relatives. Regarding MV apparatus, the mitral annulus was moderately enlarged (P<0.0001), the posterior leaflet but also the anterior leaflet were elongated (P<0.0001 and P=0.04, respectively), the posterior leaflet tip was thickened (P=0.001), and both posterior (P<0.0001) and anterior leaflets (P=0.0004) were positioned higher at end-systole in relation to the plane of mitral annulus when compared to controls. Of note, in ≈ 20% of relatives with at least a prodromal form of MVP, some degree of billowing of the anterior leaflet was associated with posterior leaflet involvement (and referred to as BiMVP in Figure 1, although leaflets did not have the classical appearance of a myxomatous form). Posterior leaflet chordal rupture was observed in 20 patients. Twenty-two patients (62±13 years) underwent MV surgery consisting of MV repair in 18 patients and MV replacement in 4 patients.

**Figure 1:**
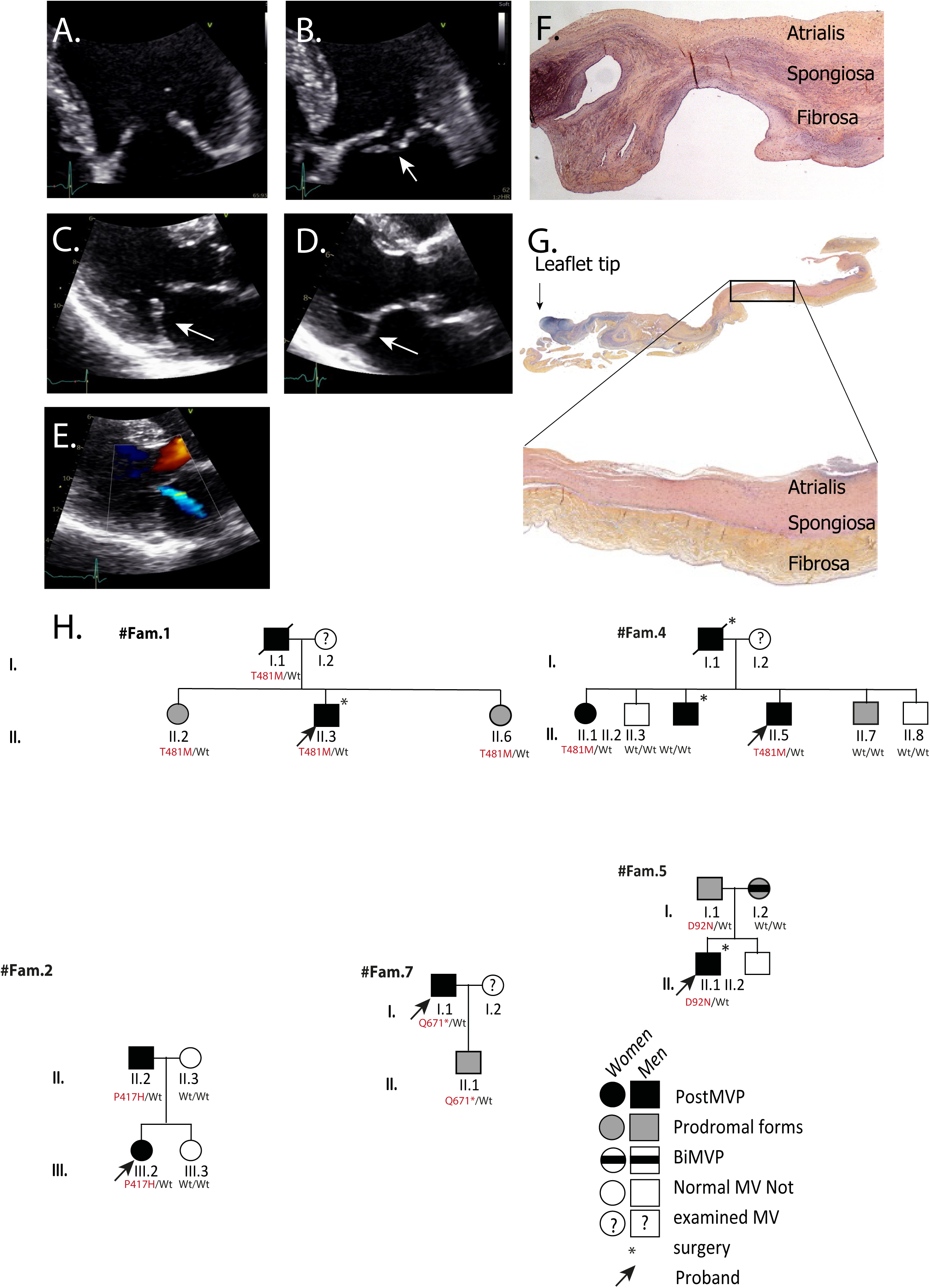
Phenotype of ARHGAP24 MVP. A and B) PostMVP dystrophy and prolapse (white arrow) with chordal rupture (flail leaflet) in diastole and systole, respectively, in a patient with *ARHGAP24* p.R95Q variant leading to mitral valve repair for severe mitral regurgitation. C, D and E) Limited dystrophy and billowing/prolapse of the posterior leaflet (white arrows) with central trace/mild mitral regurgitation in a patient with *ARHGAP24* p.T481M variant. F) Histological features of the median posterior mitral leaflet (P2) removed during surgical repair for MVP with chordae tendinae rupture in a patient with an *ARHGAP24* p.T481M variant. In the fibrosa the collagen fibers are disrupted by myxomatous infiltration of extracellular matrix. The spongiosa shows myxomatous infiltration. H&E stain; Original magnification X 2.5. During surgery leaflets were described as thin and translucent except at the level of the P2 flail scallop. G) Histological aspect of a non/minimally dystrophic part of the posterior leaflet from the same patient who underwent mitral valve replacement after a second chordae tendinae rupture. The architecture is well preserved. Proteoglycan infiltration is limited to the tip of the leaflet. The atrialis endocardium is moderately thickened. H&E stain; Original magnification X 10. H) Pedigree tree of PostMVP-familial cases carrying *ARHGAP24* variants. PostMVP patients are indicated by filled black symbols, patients with prodromal forms of MVP with gray symbols, BiMVP patients with hashed symbols, and white symbols depict individuals with normal mitral valve. The “?” sign indicates patients not examined by the clinicians. *ARHGAP24* variants carriers and non-carriers are indicated in red below symbols (Wt = wild type). Arrows show index cases. ‘*’ corresponds to patients with mitral valve surgery. *Importantly, for pre-print publication purposes, only two generations are shown at a time. Not all family members are included. The sex of relatives not relevant to the study is not displayed.*

**Table 2:** Baseline characteristics of probands with PostMVP and relatives from 20 families.

|  | All individuals | MVP patients in<br>post MVP families | Control<br>relatives | P |
| --- | --- | --- | --- | --- |
|  | n=136 | n=65 | n=71 |  |
| Age, years | 46±21 | 53±19 | 40±21 | 0.0002 |
| Age < 16 yrs, n (%) | 16 (12) | 2 (4) | 14 (20) | 0.006 |
| Male, n (%) | 69 (51) | 35 (54) | 34 (48) | 0.61 |
| Heart rate, beat/min | 68±10 | 68±9 | 69±10 | 0.71 |
| SBP, mmHg | 129±20 | 133±19 | 126±21 | 0.09 |
| AF, n (%) | 6 (5) | 6 (9) | 0 (0) | - |
| Hypertension, n (%) | 23 (17) | 17 (26) | 6 (8) | 0.01 |
| Diabetes, n (%) | 2 (1) | 1 (2) | 1 (1) | 0.52 |
| Dyslipidemia, n (%) | 19 (14) | 14 (22) | 5 (7) | 0.03 |
| Tobacco, n (%) | 42 (31) | 19 (29) | 23 (32) | 0.83 |
| MV apparatus Phenotype* |  |  |  |  |
| Mitral annulus diameter, mm | 34.0±4.8 | 35.9±5.3 | 31.9±3.2 | <0.0001 |
| AL length, mm | 22.7±3.6 | 23.4±4.3 | 22.0±2.4 | 0.04 |
| PL length, mm | 13.9±4.0 | 15.5±4.4 | 12.1±2.5 | <0.0001 |
| Tip AL thickness, mm | 2.4±0.8 | 2.4±0.8 | 2.4±0.8 | 0.92 |
| Tip PL thickness, mm | 2.4±0.8 | 2.6±0.9 | 2.1±0.6 | 0.001 |
| AL position, mm | 2.0±1.8 | 1.5±1.6 | 2.7±1.9 | 0.0004 |
| PL position, mm | -1.2±3.7 | -3.7±3.0 | 1.8±1.5 | <0.0001 |
| Anterior chordal length, mm | 23.3±4.7 | 24.1±4.7 | 22.4±4.6 | 0.09 |
| Posterior chordal length, mm | 24.7±4.7 | 25.9±5.0 | 22.8±4.0 | 0.001 |
| Chordal rupture, n (%) | 20 (15) | 20 (31) | 0 (0) | - |
| APM tip/LV length, % | 34±6 | 34±6 | 34±6 | 0.98 |
| PPM tip/LV length, % | 32±6 | 32±6 | 32±5 | 0.75 |

*Abbreviations:* MVP: mitral valve prolapse, AF: atrial fibrillation, SBP: systolic blood pressure, MV: mitral valve, AL: Anterior leaflet, PL: posterior leaflet, APM tip/LV length: anterior papillary muscle tip to left ventricular length ratio, PPM tip/LV length: posterior papillary muscle tip to left ventricular length ratio. Patients with MVP or prodromal forms are referred to as MVP patients.

### Genetic bases of inherited forms of PostMVP

To study the genetic bases of familial PostMVP, we ran WGS for the 20 PostMVP probands and selected rare variants, affecting splicing or protein amino-acids composition (analysis pipeline detailed in **Supplementary appendix**). We first screened for known genetic causes of non-syndromic MVP (*FLNA, DCHS1, DZIP1*). Three probands present with *DCHS1* variants, however, co-segregation analyses do not support causality of these variants for PostMVP (**Table S1**, **Fig. S3**) leaving PostMVP familial cases free of known genetic origin of inherited MVP.

We thus searched for potential novel genetic origins/risk factors for MVP in the families by performing a burden analysis, which consists in searching for genes statistically enriched in rare variants in PostMVP probands (n=20) when compared to a large origin-matched control population (n=856) (**Fig. S1** and **supplementary appendix**). We identified an enrichment (p-value <0.001) in three genes with at least 4 mutated patients (*IFT140 (*p = 9.55e-07), *ARHGAP24 (*p = 3.06e-04) and *ZNF221 (*p = 6.2e-04)) (**Table S2**).

*ARHGAP24* encodes the Filamin A binding RhoGTPase-activating protein (FilGAP). It appears as a highly relevant MVP related gene as it interacts with Filamin A (FlnA), the first identified causal gene responsible for familial forms of MVP^8,12^. Based on the clinical and fundamental knowhow of the team acquired on the investigations on *FLNA*-related phenotypes and pathways^8,12,21,29^, we focused our investigations on the phenotype of *ARHGAP24-*mutated probands and relatives in regards to PostMVP, evaluate the effects of the silencing of ARHGAP24 *in vivo* and test the effects of identified variants on the natural function of FilGAP *in vitro*.

### Phenotype of *ARHGAP24* MVP

By running co-segregation analyses of *ARHGAP24* variants (namely *ARHGAP24* p.D92N, p.P417H, p.T481M, p.Q671*) with the PostMVP phenotypes in five families, we found that among 11 PostMVP patients, 10 carry the *ARHGAP24* variants (**Fig. 1H, Tab. S1**). Also, of 9 patients with prodromal forms 5 carry *ARHGAP24* variants. Three relatives, mostly young patients (Fam.1: III:2, IV:5 and Fam.2: IV:1), carry the identified variants and present with a normal valve. Of note, two independent families (Fam.1 and 4 (**Fig.1H**)) carry the same variant *ARHGAP24-*p.T481M. **Fig. 1A to G** illustrate the phenotype of *ARHGAP24*-related PostMVP. In the absence of chordal rupture, mitral valve dystrophy of the posterior leaflet was limited to billowing/discrete prolapse and elongation with only trace/mild central MR. Chordal rupture leads to the typical aspect of flail leaflet with severe MR encountered in particular in FED type MVP.

Mitral valve apparatus quantitative assessment in 16 adults (>16 years old, 52±18 years) carrying *ARHGAP24* variants was compared with 54 healthy adult relatives (48±16 years, P=0.31). Patients with *ARHGAP24* variants had enlarged mitral annulus diameter (36.4±5.1 vs. 31.7±3.0 mm, P<0.0001), significant elongation (16.4±4.4 vs. 11.9±2.5 mm, P<0.0001) and billowing/prolapse (−2.9±2.2 vs. 1.9±1.6 mm, P<0.0001) of the posterior mitral valve leaflet consistent with the phenotype of PostMVP in the overall population.

### Genetic screening of *ARHGAP24* in the 284 MVP cases

In order to evaluate the proportion of *ARHGAP24* genetic variations in a broader MVP population, we screened *ARHGAP24* in the 264 other MVP unrelated patients (86 PostMVP patients and 178 BiMVP patients) (**Fig. S1**). We found 8 additional variant carriers, 3 PostMVP patients (p.R95Q, p.T481M and p.M680L) and 5 BiMVP patients (**Table S3**). Overall, 13 out of 284 patients carried rare *ARHGAP24* variants (**Fig. S4**). We observed a significant enrichment in rare *ARHGAP24* variants in PostMVP cases compared to controls (8 PostMVP carriers out of 106 PostMVP patients *vs.* 24 carriers out of 856 controls, p=0.018). There was no significant enrichment for BiMVP (5 BiMVP carriers out of 178 BiMVP patients, p=1.0).

### Silencing of ARHGAP24 *in vivo* and atrioventricular valve dysfunction

To evaluate the role of *ARHGAP24* in the mitral valve function, we first knocked down the expression of the zebrafish ortholog (ENSDARG00000060175) with an antisense morpholino oligonucleotide (**Fig. S5A and B** (∼25% reduction)). The resulting FilGAP knockdown embryos were morphologically normal with unperturbed body plan, axis and no change in mortality rate (**Fig. 2A and Fig. S5C**). To evaluate valvular development and function, we assayed the percentage of embryos displaying atrioventricular valve regurgitation at 96 hours post fertilization by high-speed videography. FilGAP knockdown resulted in a significant increase in the proportion of embryos with atrioventricular valve regurgitation (41/157, 26.1%, P=0.004) when compared to control embryos receiving non-targeting morpholino (15/117, 12.8%,) (**Fig. 2B and C, Suppl. video data**). These results were replicated with a second, independent morpholino (30/107, 28.0%, P=0.0004). As gross cardiac chamber morphology and looping was unperturbed (**Fig. 2A and C**), together, these data suggest a specific and critical role for FilGAP in the function of the atrioventricular valve as observed in *ARHGAP24* variants carriers.

**Figure 2:**
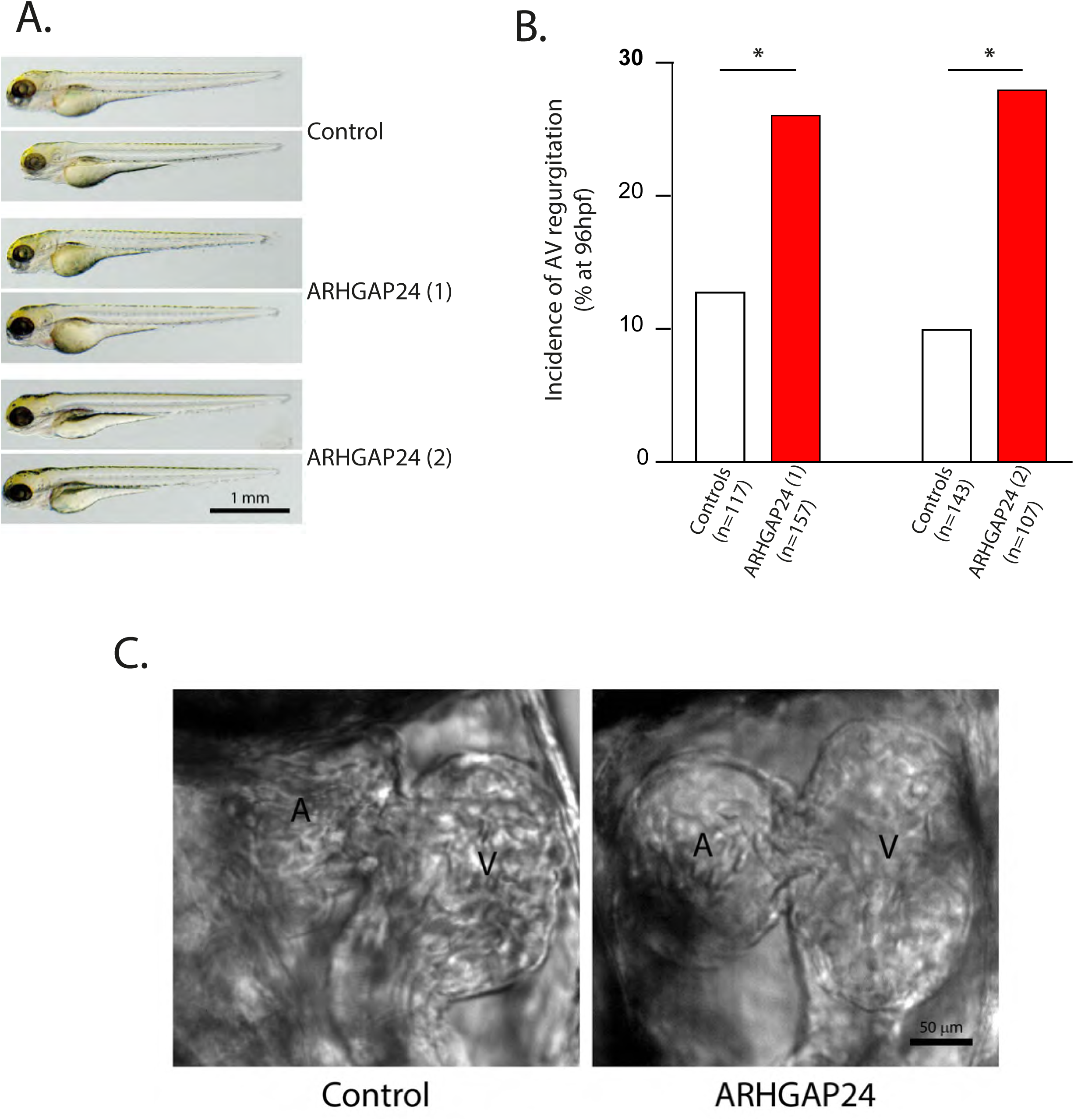
Knockdown of the ortholog of ARHGAP24 induces atrio-ventricular regurgitation in zebrafish. A) ARHGAP24 knockdown using two specific morpholinos (ARHGAP24, 1 and 2) does not affect morphological development nor cardiac chamber morphology, B) but significantly increased atrio-ventricular blood regurgitation (C). * Indicates significant statistical difference (p<0.05) with control morpholino. Representative videos of blood regurgitation are available as **supplemental materials**.

### *In vitro* assessment of genetic *ARHGAP24* variants impact on FilGAP functions

While it appears moderately expressed in the heart (**Fig. S6A**), FilGAP is highly expressed in human mitral valves (compared to brain, kidney and total heart, **Fig. S6B**). More specifically, the FilGAP protein is located in both human valvular endothelial and interstitial cells (**Fig. S6C**). At the cellular level, FilGAP is known to modulate cellular spreading and adhesion mechanisms in which it functions as a dimer ^19,30,31^. Futhermore, as a natural inhibitor of the Rac1-GTPase, FilGAP is involved in the remodeling of the actin cytoskeleton, the regulation of cellular adhesion processes and mechanical cell properties (**Fig. 3A**).

**Figure 3:**
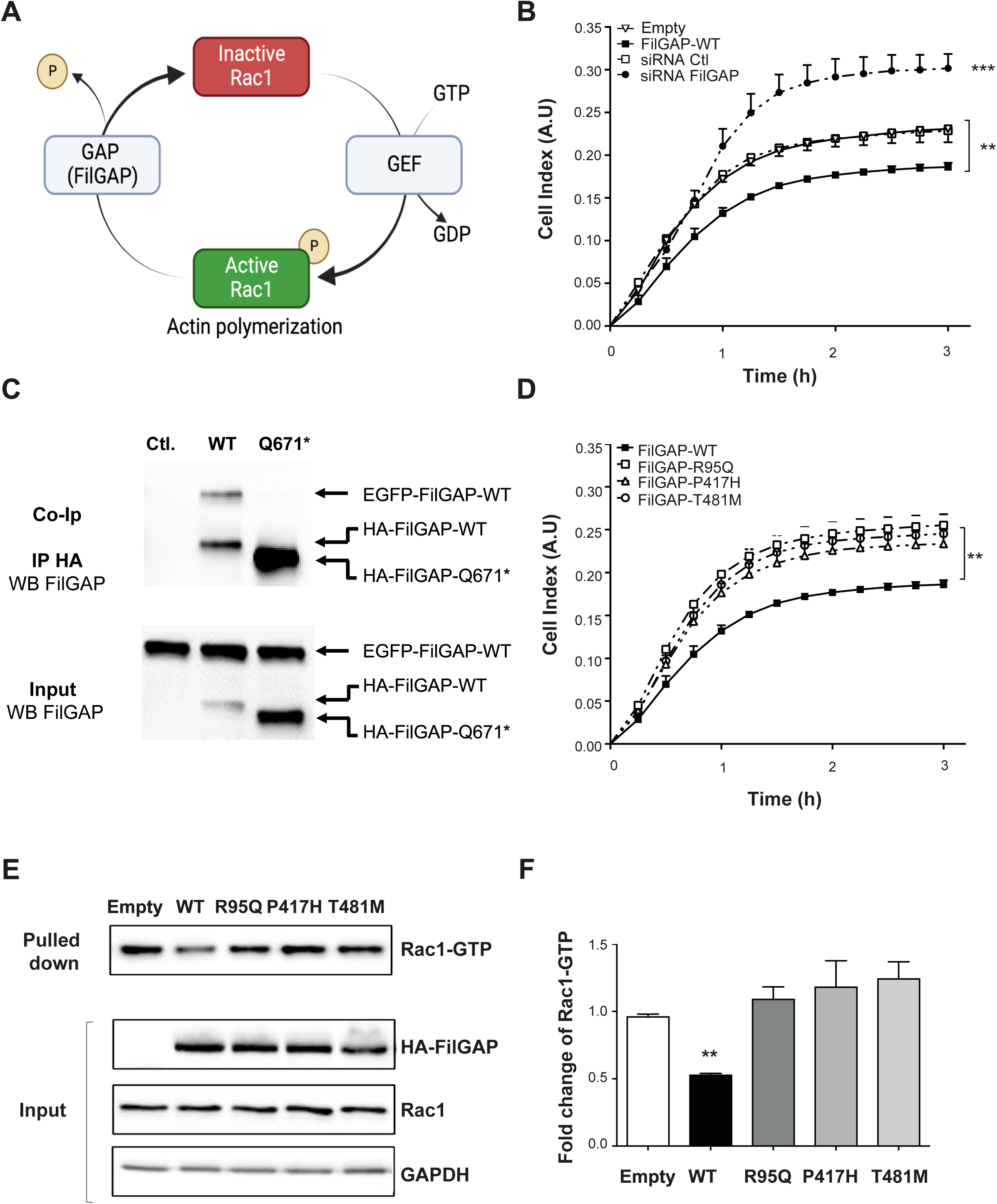
*ARHGAP24* PostMVP-associated variants impact the function of the protein (FilGAP, ENSP00000378611) A) Activation/Inactivation cycle of Rac1 mediated by GTPase-activating protein (GAP) and guanine nucleotide exchange factors (GEF). B) Cell indexes (CI) values measured using xCELLigence system. FilGAP-WT transfected cells exhibited slower CI increases and lower steady states CI (CIst) compared to pcdna3 or siRNA control treated cells. On the other hand, cells treated with siRNA targeting FilGAP exhibited faster increase and higher CIst compared to controls. C) EGFP-tagged FilGAP-WT was co-immunoprecipitated with the HA-tagged FilGAP WT but not Q671* mutant. Hek293T cells were transfected with EGFP-tagged FilGAP-WT only (ctl.); or co-transfected with HA-tagged FilGAP (WT or Q671*. HA-tagged FilGAPs were immunoprecipitated with anti-HA antibody and blots revealed using anti FilGAP antibody. The upper panel (Co-IP) shows the co-immunoprecipitated fraction and the lower panel (input) the total protein fraction. D) Cell indexes (CI) recordings obtained from R95Q, P417H and T481M transfected cells. None of the mutant FilGAPs altered cell spreading and adhesion compared to pcdna3 transfected cells. Values represent the mean of six experiments and SEM are indicated by the bars. ** significant difference (p<0.01) *vs.* FilGAP-WT. E) Typical western blot of a Rac1-GTP pull down assay experiment. Hek293T cells were transfected with pcdna3 (control), HA-FilGAP-WT, R95Q, P417H or T481M and active Rac1 isolated using GST-PAK1. Although all HA-FilGAP constructs were expressed to similar levels (input FilGAPs-HA) only FilGAP-WT (second lane) significantly decreased the active Rac1 pulled down compared to control. F) The histogram on the right gives the mean data from seven experiments. Error bars show SEM, ** P<0.01 versus pcdna3 condition. ** indicates significant difference (P<0.01) versus FilGAP-WT cells.

By monitoring cell adhesion and spreading we first show, as previously demonstrated^30,31^, that the silencing of *ARHGAP24* with si-RNA increases cell adhesion capacities (p<0.001 compared to control) (**Fig. S7, Fig. 3B**). Inversely, its overexpression reduced cell adhesion compared to controls (p<0.01) (**Fig. 3B**).

#### ARHGAP24 variants impair cellular adhesion and spreading capacities of cells

As FilGAP functions as dimers, we secondly tested the ability of mutated FilGAP to dimerize with WT-FilGAP intracellularly. Using co-immunoprecipitations, we found that only p.Gln671Ter, which induces a premature truncation of the protein affects FilGAP dimerization (**Fig. 3C**). The others variants did not impact the dimerization process (*data not shown*).

To evaluate the functional impact of *ARHGAP24* variants identified, we further tested the adhesion and spreading capacities of cells transfected with WT or mutated forms of FilGAP (p.R95Q, p.P417H and p.T481M). These mutations significantly impaired cellular adhesion and spreading capacities (p<0.01, **Fig. 3D**).

Taken together, these data suggest that the mutations identified induce a loss of function of FilGAP. Of note, no effect was observed for the *ARHGAP24-*p.D92N variant (*data not shown*).

#### Mutated-FilGAP forms lose their GTPase-activating capacities

Based on previous findings showing that the variants *ARHGAP24*-p.R95Q, -p.P417H and - p.T481M impaired cellular adhesion and spreading capacities, we hypothesized that it might be due to a loss of function on FilGAP-GTPases’ activity on Rac1.

As shown on **Fig. 3E and F**, the overexpression of WT-FilGAP decreases the amount of activated Rac1 (bound to GTP) compared to controls (P<0.01), while it remains unchanged in FilGAP mutated conditions (R95Q, P417H and T481M). No effects were found for p.D92N variants (*data not shown*). Together these data indicated that the three mutations identified (R95Q, P417H, T481M) affect the GTPase activating protein (GAP) capacity of FilGAP on Rac1 and subsequently cellular adhesion and spreading capacities.

## DISCUSSION

We were able to identify and describe for the first-time inherited forms of PostMVP in 20 families. The recurrence of both typical and prodromal MVP forms was around 40% in relatives of PostMVP probands. Although alteration of the mitral valve apparatus predominates on the posterior leaflet in relatives, some degree of anterior leaflet dystrophy has also been demonstrated.

Genetic analyses, using burden testing, found that rare variants in *ARHGAP24* are risk factors associated with PostMVP, with an association between *ARHGAP24* variants and typical or prodromal PostMVP in families. We subsequently demonstrated that the silencing of *ARHGAP24* in zebrafish affects the functionality of the atrioventricular valve. Finally, *in vitro* analyses showed that identified genetic variants are loss-of-function variants which impair cellular adhesion and mechano-transduction capacities.

### PostMVP inheritance

PostMVP, also called FED, is considered to exhibit a different phenotype compared to myxomatous BiMVP on both macroscopic and histologic examination^4,32^. PostMVP is generally characterized by thin translucent leaflets, localized dystrophy of a single leaflet segment, mild to moderate mitral annulus dilatation, thin chordae tendineae that can rupture spontaneously especially at the posterior leaflet, resulting in flail leaflet and acute severe MR. Although MV apparatus is generally moderately altered in PostMVP, we found significant differences in PostMVP patients compared to control relatives regarding mainly posterior leaflet, mitral annulus, and chordae tendinae, but also minor phenotypic modifications of the anterior leaflet length and position generally not detectable on a qualitative approach. Taken together, these findings suggest that the pathophysiological processes involved in PostMVP lead to a discrete but global alteration of the MV apparatus. Further, some degree of anterior leaflet billowing were observed in ≈20% of relatives with posterior leaflet involvement (and referred to BiMVP in Figure 1). This may be explained either by a stronger expression of the disease in some individuals or by the interaction of different genetic backgrounds in the same family, in agreement with both monogenic and polygenic background of MVP according to recent publications^5,8,13–15,17^. In fact, the analysis of putative monogenic BiMVP myxomatous families has shown a wide variety of mitral valve involvement, from bi-leaflet prolapse to monoleaflet prodromal forms^6,8^. Whatever the explanation, this finding may suggest that the separation between PostMVP and BiMVP is blurred, and that there is, at least on an echocardiographic approach, a continuum of MVP expression from forms with localized dystrophy (FED+), to moderately (Barlow fruste) or severely dystrophic and myxomatous (Barlow) forms^32^.

Previous publications demonstrated the familial aggregation of classical myxomatous BiMVP ^8,11,33,34^. The mode of inheritance is predominantly autosomal dominant except in the X-linked FLNA-MVP case^12^. By contrast, PostMVP was usually described as a pure degenerative age-related disorder and familial aggregation was not reported but likely not investigated. The recurrence of MVP in PostMVP families, including prodromal forms, was around 40%. This observation constitutes, to our knowledge, the first report of inherited forms of PostMVP. This finding strongly suggests a genetic origin of PostMVP. We could however not expand these results to flail leaflets occurring in very old patients without any evidence of leaflet segment dystrophy. In fact, these patients were separately stratified in the classification proposed by Adams et al.^32^.

### Genetic risk factors for PostMVP

Noteworthy, the top associated genes, *ARHGAP24* and *IFT140* are involved in mechanosensing and transduction, two major MVP-related pathways.

*ARHGAP24,* which encodes the Filamin A binding RhoGTPase-activating protein (FilGAP), appears as a relevant MVP related gene as it interacts with Filamin A (FlnA), the first identified causal gene of MVP^8,12^. In addition, Gould et al. previously showed that FilGAP is involved in the coordination of heart valve morphogenesis during development via its action on small GTPases (Rac1 and RhoA more specifically) in chicks^35^. *IFT140*, which stands for intra-flagellar transport protein 140, encodes one of the subunits of the intra-flagellar transport (IFT) system (complex-A) involved in the signaling of primary cilia, a key mechanism involved in MVP pathogenesis^13,15,36^. However, additional functional investigations are needed to confirm that *IFT140* can be involved in the pathophysiology of PostMVP.

While the top associated genes, *IFT140* and *ARHGAP24*, are involved in major MVP-related pathways, the functional involvement of *ZNF221* remains more hypothetical and will need further investigations.

### Clinical presentation of *ARHGAP24* mutation carriers

On the clinical point of view*, ARHGAP24* variant carriers present with an enlarged mitral annulus diameter, an increased billowing of the posterior leaflet and an elongated posterior mitral valve leaflet. Coherent with the late onset of PostMVP, the youngest mutation carriers did not exhibit mitral valve dysfunction while the oldest presented clear PostMVP with eventual MR related to chordal rupture supporting the age-related degenerative features of the pathology (**Fig. 1**). While FilGAP is a well-known partner of Filamin A, notable phenotypic differences exist between FlnA and FilGAP mutated patients. FlnA MVP-patients exhibit polyvalvular defects associated with developmental and degenerative myxomatous alterations of both anterior and posterior mitral leaflets as well as of the sub valvular apparatus but no chordal rupture. Such a clinical portrait is more reminiscent of Barlow’s disease as opposed to PostMVP studied here^8^. FilGAP-associated mitral valve disorders are, in contrast, restricted to the mitral valve apparatus and especially the posterior leaflet when compared to FlnA, with a later development of mitral valve disease in life. Previous studies by Akilesh et al. linked the *ARHGAP24* mutations to focal segmental glomerulosclerosis ^37^. However, in the present study, none of the PostMVP patients exhibited any sign of renal dysfunction.

### FilGAP, valve development and molecular mechanisms involved

We showed that morpholino mediated knockdown of FilGAP in the Zebrafish resulted in atrioventricular regurgitation. This observation is in line with the study from Gould et al. who showed that FilGAP is involved in the Rac-1 mediated fetal-valve remodeling in chicks^35^. Together, these data point to the small GTPases pathway and their role in the mechano-transduction pathways as critical factors of heart valve development and homeostasis^19,21,38^.

The FilGAP/FlnA complex, acting on actin-cytoskeleton integrity and polymerization appears as a key actor involved in MVP pathogenesis. Consistent with the role of the FilGAP/FlnA complex in the protection of cells submitted to mechanical stress, Shifrin et al. showed that FilGAP localization changes as it is recruited to force transfer sites when cells are submitted to mechanical constraints ^39^. One can speculate that the mutant FilGAPs may weaken interactions with FlnA and subsequently affect its recruitment to these sites. This mis-localization might inhibit Rac1-activity or make it accessible to regulatory factors including the RhoA activated kinase ROCK or Arf6 ^40,41^ with the potential involvment of the periostin/integrin-β1 pathway^42^. Consistent with this idea, mice deficient in the endothelial Rac1 activator Dock180 exhibit cardiac abnormalities including valve malformations. Future studies will be necessary to clarify these points. Overall, genetic and functional analyses show that *ARHGAP24* variants gather the characteristics of rare variants with strong effect in PostMVP pedigrees.

### Limitations

We have to acknowledge some limitations in our study, mainly due to the limited number of patients carrying *ARHGAP24* variants. Despite small effectives, these patients present with a quite homogeneous PostMVP phenotype which supports the validity of our genetic unbiased findings. Also, in some familial cases we observed incomplete penetrance of *ARHGAP24* variants with the PostMVP phenotype. This observation is in line with previous familial and pathophysiological studies which described a complex pattern of inheritance and familial expression of MVP. These genotype-phenotype discrepancies, commonly observed in genetic investigations of MVP-familial cases, are also likely due to the late-onset development of the disease as well as its relatively high frequency in the general population. The screening of broader populations and larger MVP-pedigrees will reinforce the causative role of *ARHGAP24* as an MVP gene. The present study does not address the question of why the posterior leaflet is predominantly involved. One can speculate that, as the two leaflets present with embryologically different origins, differential cellular responses might be compounded by differences in leaflet and/or chordal stress.

## Conclusion

The present study shows that the PostMVP phenotype, often referred as fibroelastic deficiency, can be inherited and have specific genetic and pathophysiological origins. Moreover, with the identification of *ARHGAP24* as a susceptibility gene for MVP, we reinforce the evidence pointing at the pivotal role of the mechano-transduction pathways in the pathogenesis of MVP.

## Data Availability

All data produced in the present work are contained in the manuscript.

## ACKNOWLEDGEMENTS

We thank Marie Marrec and Guénola Coste for their assistance in patients’ recruitment and family screening, as well as the biological resource centre for biobanking (CHU Nantes, CRB, BRIF: BB-0033-00040). We are most grateful to the Bioinformatics Core Facility of Nantes BiRD, member of Biogenouest, Institut Français de Bioinformatique (ANR-11-INBS-0013), for the use of its resources and for its technical support. We thank the IBISA MicroPICell facility (Biogenouest), a member of the national infrastructure France-Bioimaging (ANR-10-INBS-04), as well as the Therassay and the UTE facilities of the Structure Fédérative de Recherche François Bonamy (Nantes, France) for technical assistance.

This work was funded by The Leducq Foundation, Transatlantic Mitral Network of Excellence grant 07CVD04 (Paris, France), the Clinical Research Hospital Program (PHRC) from the French Ministry of Health PHRC-I (2012), the “Fédération Française de Cardiologie” (2011) (TLT, JM) (FFC, Paris, France), the “Fondation Coeur et Recherche” 2015 (TLT), and the French Ministry of Research: ANR grant ANR-16-CE17–0015-01 (to J.-J.S. and N.B.-N). The work at MUSC was performed in a facility constructed with support from the National Institutes of Health, Grant Number C06 RR018823 from the Extramural Research Facilities Program of the National Center for Research Resources. Other funding sources: National Heart Lung and Blood Institute: R01-HL33756 (RRM), COBRE 1P30 GM103342 (RRM, RAN), 8P20 GM103444 (RRM and RAN), R01-HL127692 (RAN); American Heart Association: 11SDG5270006 (RAN); National Science Foundation: EPS-0903795 (RRM); NHLBI K24, HL67434 and HL109506 (RAL). The sequencing work was funded by the France Génomique National infrastructure, funded as part of the « Investissements d’Avenir » program managed by the Agence Nationale pour la Recherche (contract ANR-10-INBS-09). RC was supported by the the European Union’s Horizon 2020 research and innovation program under the Marie Sklodowska-Curie grant agreement No. 846291, by the French Society of Cardiology under the “Alain Castaigne” scientific prize and by a “Connect Talent” research chair from Région Pays de la Loire and Nantes Métropole. AR was supported by postdoctoral fellowship grants from the Institut de France–Fondation Lefoulon-Delalande and from the ‘Fondation pour la Recherche Médicale’ (FRM: EQU201903007846, project INSTINCTIVE).

---

**SUPPLEMENTAL FIGURE 1.**
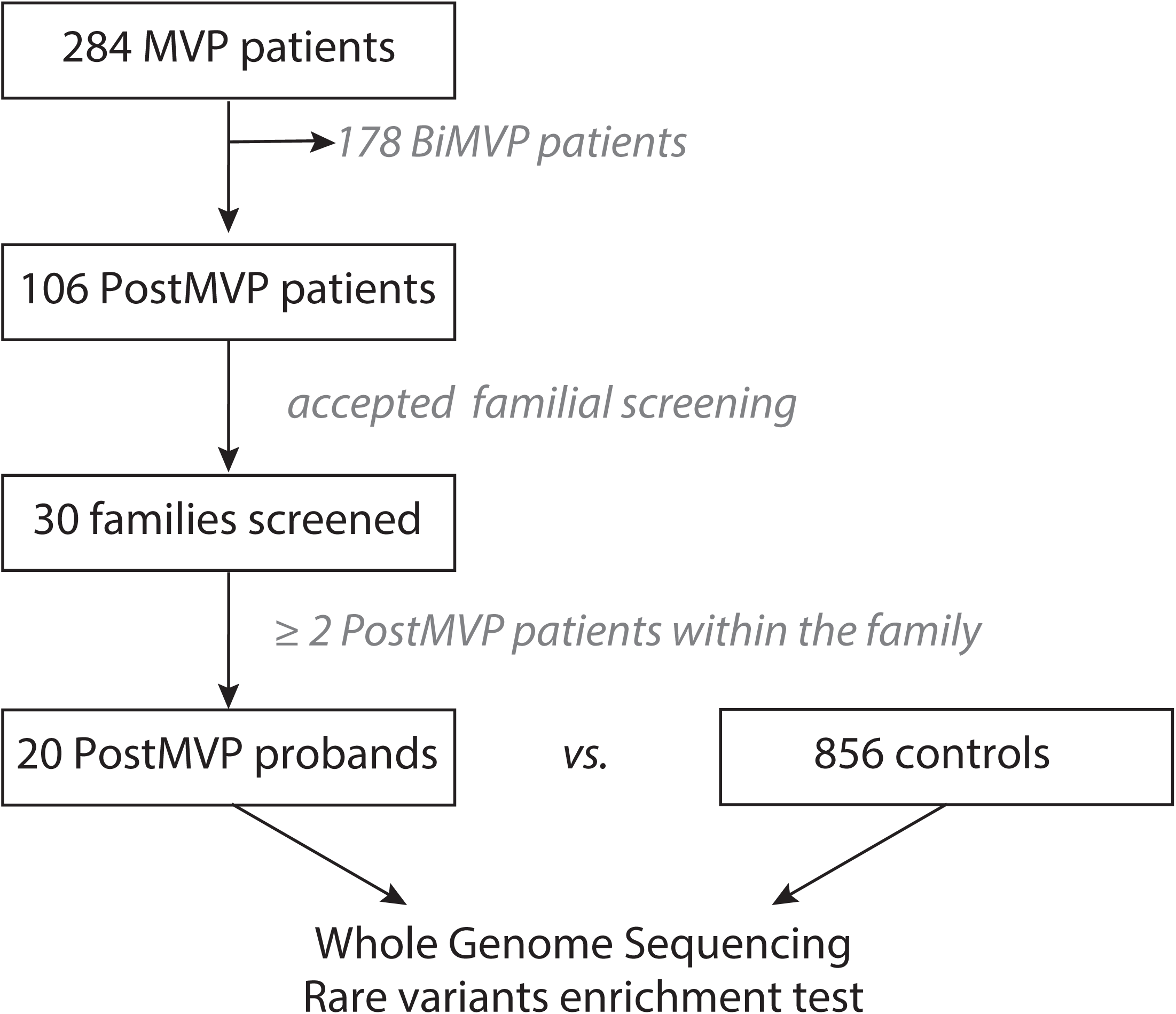

**SUPPLEMENTAL FIGURE 2.**
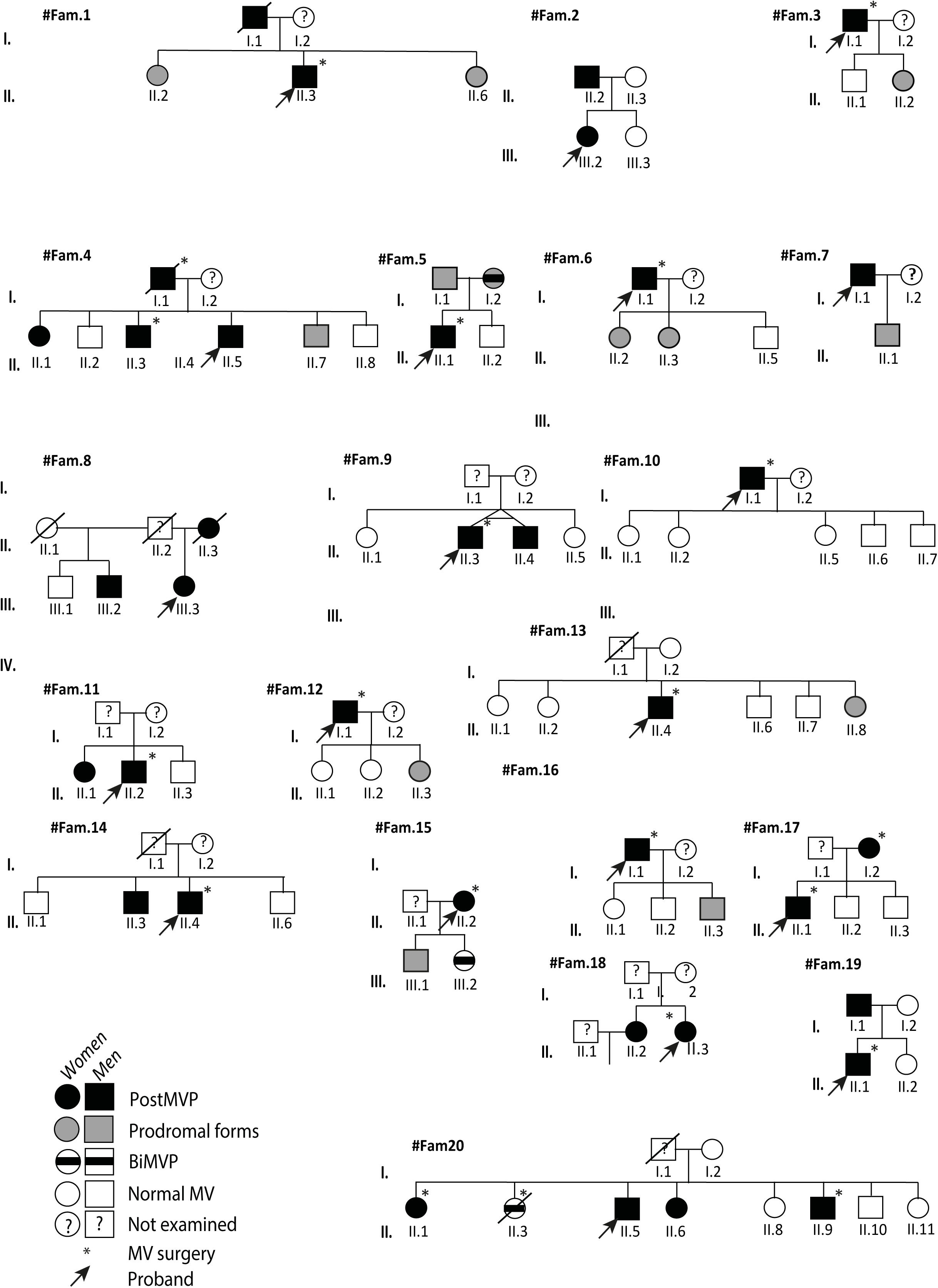

**SUPPLEMENTAL FIGURE 3.**
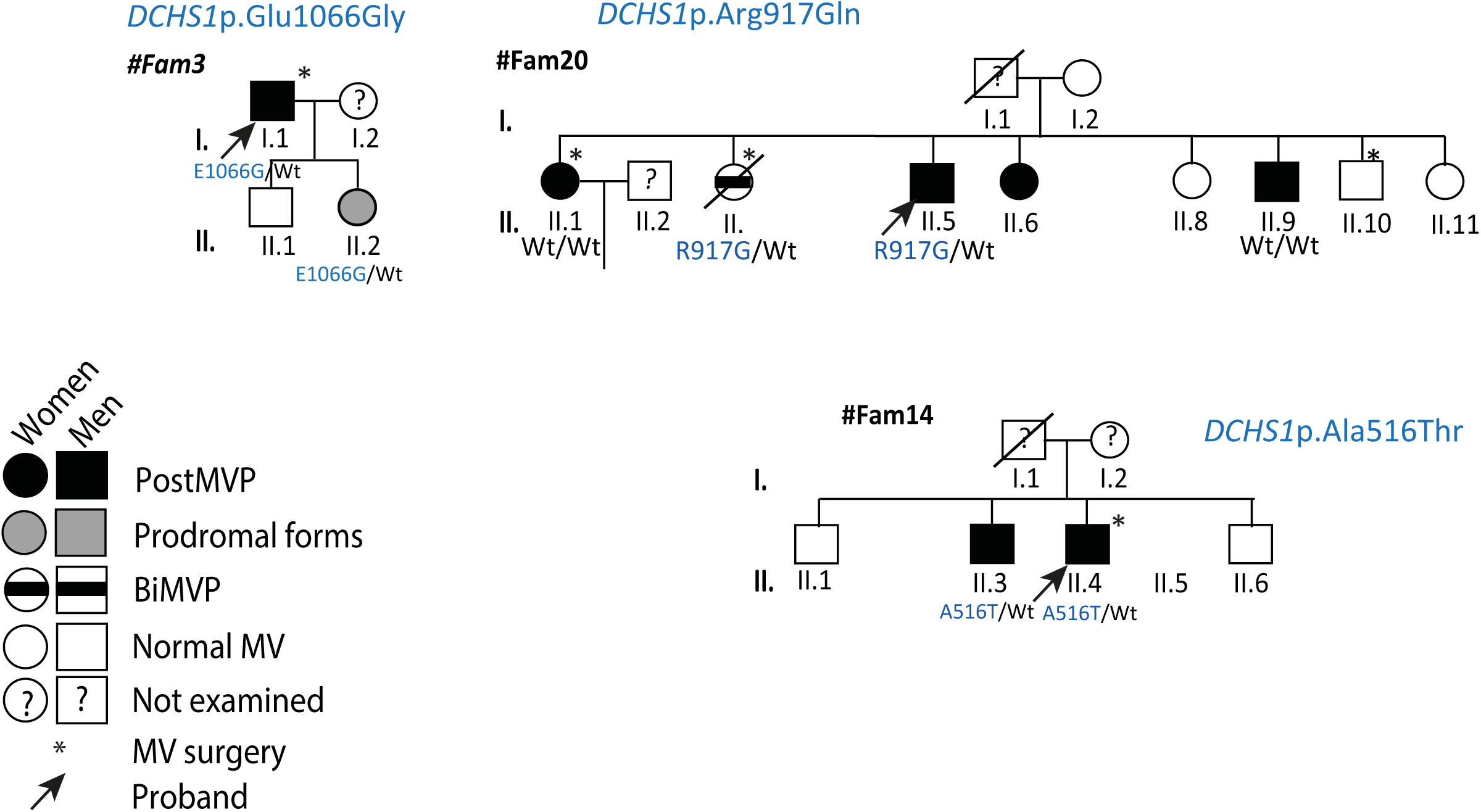

**SUPPLEMENTAL FIGURE 4.**
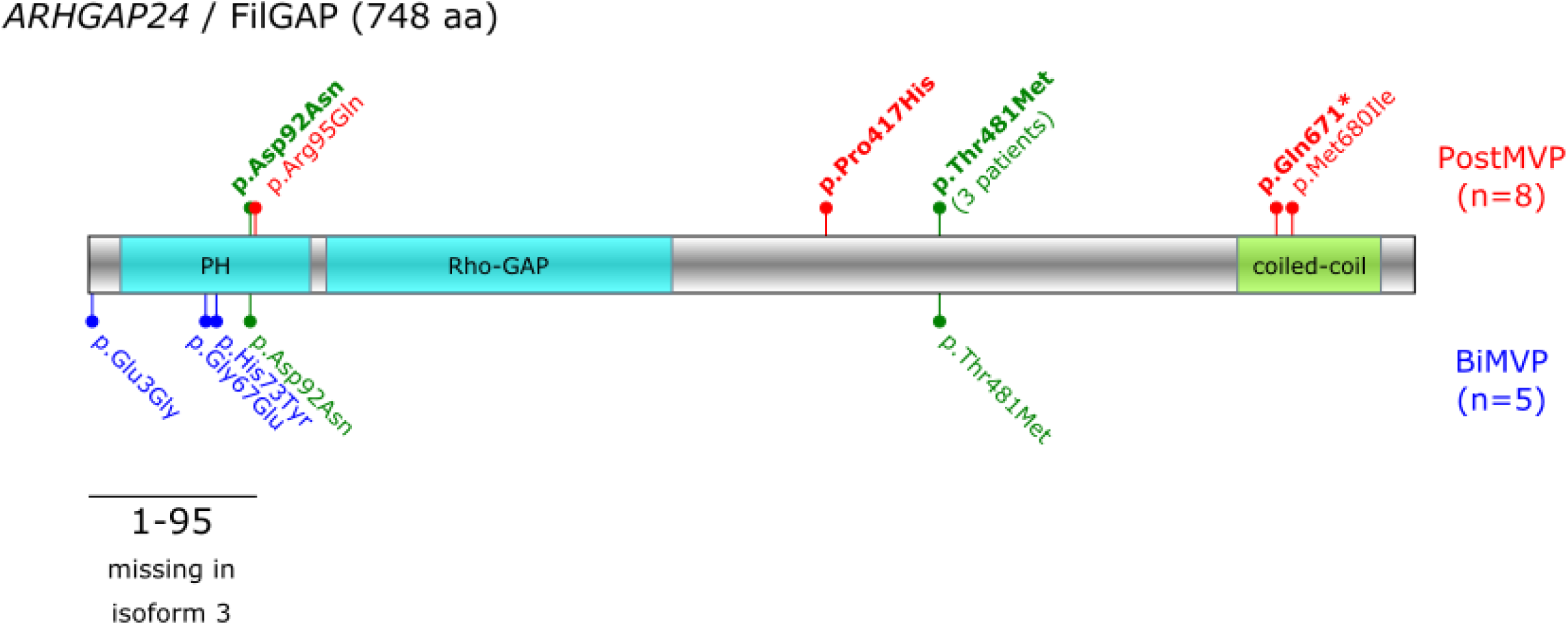

**SUPPLEMENTAL FIGURE 5.**
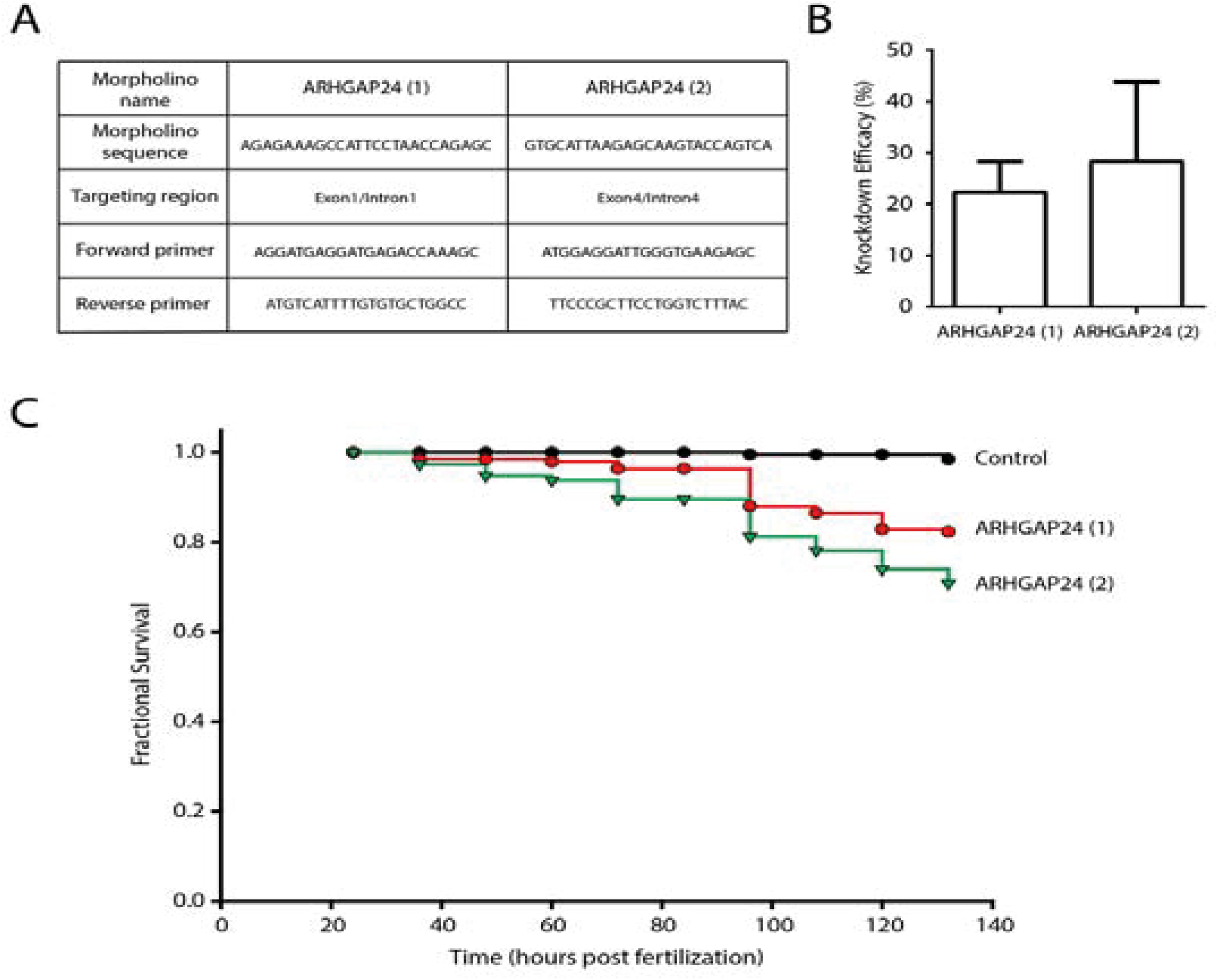

**SUPPLEMENTAL FIGURE 6.**
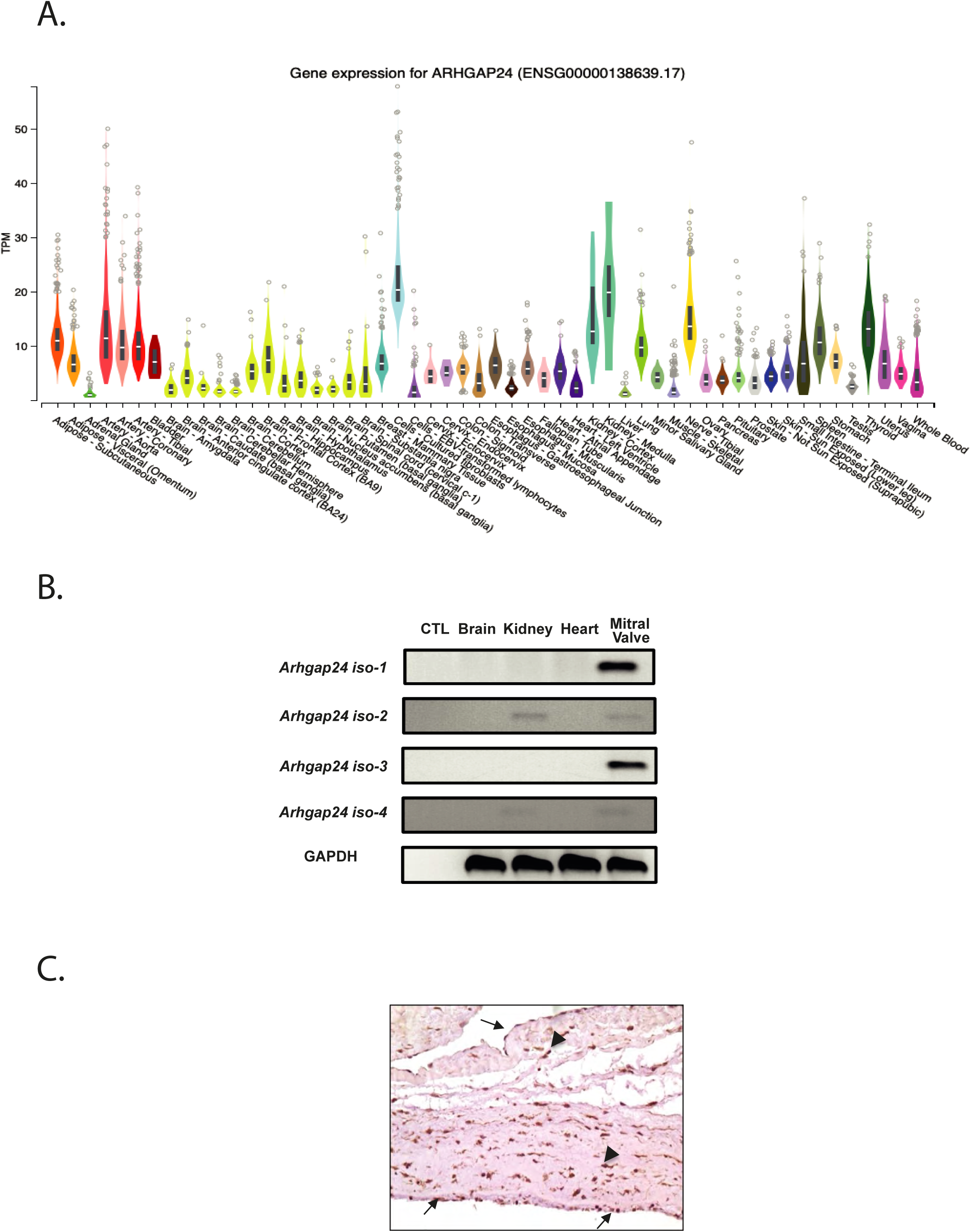

**SUPPLEMENTAL FIGURE 7.**
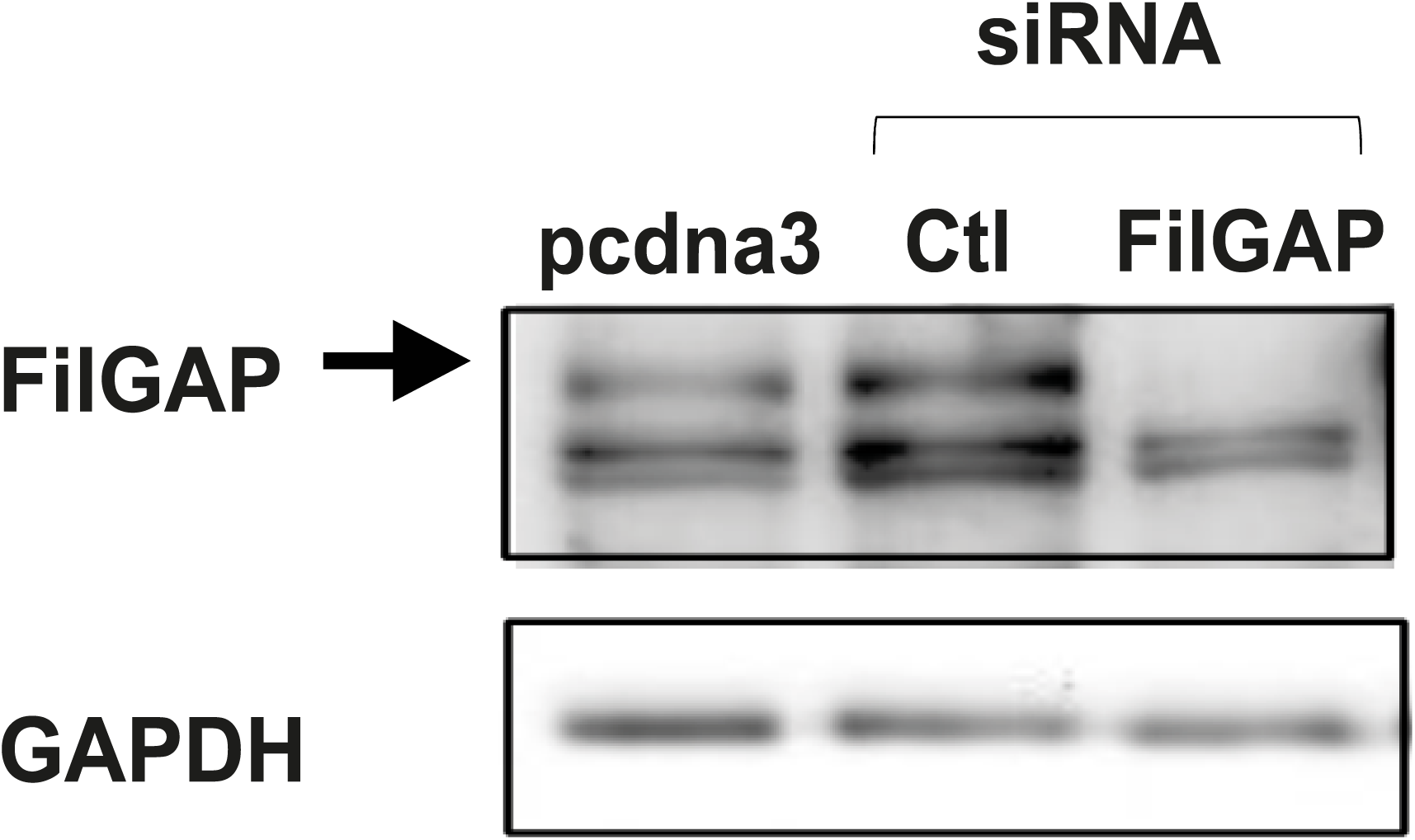

## REFERENCES

1. Levine RA, Hagége AA, Judge DP, Padala M, Dal-Bianco JP, Aikawa E, Beaudoin J, Bischoff J, Bouatia-Naji N, Bruneval P, Butcher JT, Carpentier A, Chaput M, Chester AH, Clusel C, Delling FN, Dietz HC, Dina C, Durst R, Fernandez-Friera L, Handschumacher MD, Jensen MO, Jeunemaitre XP, Le Marec H, Le Tourneau T, Markwald RR, Mérot J, Messas E, Milan DP, Neri T, Norris RA, Peal D, Perrocheau M, Probst V, Pucéat M, Rosenthal N, Solis J, Schott J-J, Schwammenthal E, Slaugenhaupt SA, Song J-K, Yacoub MH, Leducq Mitral Transatlantic Network. Mitral valve disease--morphology and mechanisms. Nat Rev Cardiol. 2015;12:689–710.

2. Freed LA, Levy D, Levine RA, Larson MG, Evans JC, Fuller DL, Lehman B, Benjamin EJ. Prevalence and clinical outcome of mitral-valve prolapse. N Engl J Med. 1999;341:1–7.

3. Delling FN, Rong J, Larson MG, Lehman B, Osypiuk E, Stantchev P, Slaugenhaupt SA, Benjamin EJ, Levine RA, Vasan RS. Familial clustering of mitral valve prolapse in the community. Circulation. 2015;131:263–268.

4. Fornes P, Heudes D, Fuzellier JF, Tixier D, Bruneval P, Carpentier A. Correlation between clinical and histologic patterns of degenerative mitral valve insufficiency: a histomorphometric study of 130 excised segments. Cardiovasc Pathol. 1999;8:81–92.

5. Le Tourneau T, Mérot J, Rimbert A, Le Scouarnec S, Probst V, Le Marec H, Levine RA, Schott J-J. Genetics of syndromic and non-syndromic mitral valve prolapse. Heart. 2018;104:978–984.

6. Nesta F, Leyne M, Yosefy C, Simpson C, Dai D, Marshall JE, Hung J, Slaugenhaupt SA, Levine RA. New locus for autosomal dominant mitral valve prolapse on chromosome 13: clinical insights from genetic studies. Circulation. 2005;112:2022–2030.

7. Delling FN, Rong J, Larson MG, Lehman B, Fuller D, Osypiuk E, Stantchev P, Hackman B, Manning WJ, Benjamin EJ, Levine RA, Vasan RS. Evolution of Mitral Valve Prolapse: Insights From the Framingham Heart Study. Circulation. 2016;133:1688–1695.

8. Le Tourneau T, Le Scouarnec S, Cueff C, Bernstein D, Aalberts JJJ, Lecointe S, Mérot J, Bernstein JA, Oomen T, Dina C, Karakachoff M, Desal H, Al Habash O, Delling FN, Capoulade R, Suurmeijer AJH, Milan D, Norris RA, Markwald R, Aikawa E, Slaugenhaupt SA, Jeunemaitre X, Hagège A, Roussel J-C, Trochu J-N, Levine RA, Kyndt F, Probst V, Le Marec H, Schott J-J. New insights into mitral valve dystrophy: a Filamin-A genotype-phenotype and outcome study. Eur Heart J. 2018;39:1269–1277.

9. Monteleone PL, Fagan LF. Possible X-linked congenital heart disease. Circulation. 1969;39:611–614.

10. Kyndt F, Schott JJ, Trochu JN, Baranger F, Herbert O, Scott V, Fressinaud E, David A, Moisan JP, Bouhour JB, Le Marec H, Bénichou B. Mapping of X-linked myxomatous valvular dystrophy to chromosome Xq28. Am J Hum Genet. 1998;62:627–632.

11. Devereux RB, Brown WT, Kramer-Fox R, Sachs I. Inheritance of mitral valve prolapse: effect of age and sex on gene expression. Ann Intern Med. 1982;97:826–832.

12. Kyndt F, Gueffet J-P, Probst V, Jaafar P, Legendre A, Le Bouffant F, Toquet C, Roy E, McGregor L, Lynch SA, Newbury-Ecob R, Tran V, Young I, Trochu J-N, Le Marec H, Schott J-J. Mutations in the gene encoding filamin A as a cause for familial cardiac valvular dystrophy. Circulation. 2007;115:40–49.

13. Durst R, Sauls K, Peal DS, deVlaming A, Toomer K, Leyne M, Salani M, Talkowski ME, Brand H, Perrocheau M, Simpson C, Jett C, Stone MR, Charles F, Chiang C, Lynch SN, Bouatia-Naji N, Delling FN, Freed LA, Tribouilloy C, Le Tourneau T, LeMarec H, Fernandez-Friera L, Solis J, Trujillano D, Ossowski S, Estivill X, Dina C, Bruneval P, Chester A, Schott J-J, Irvine KD, Mao Y, Wessels A, Motiwala T, Puceat M, Tsukasaki Y, Menick DR, Kasiganesan H, Nie X, Broome A-M, Williams K, Johnson A, Markwald RR, Jeunemaitre X, Hagege A, Levine RA, Milan DJ, Norris RA, Slaugenhaupt SA. Mutations in DCHS1 cause mitral valve prolapse. Nature. 2015;525:109–113.

14. Dina C, Bouatia-Naji N, Tucker N, Delling FN, Toomer K, Durst R, Perrocheau M, Fernandez-Friera L, Solis J, PROMESA investigators, Le Tourneau T, Chen M-H, Probst V, Bosse Y, Pibarot P, Zelenika D, Lathrop M, Hercberg S, Roussel R, Benjamin EJ, Bonnet F, Lo SH, Dolmatova E, Simonet F, Lecointe S, Kyndt F, Redon R, Le Marec H, Froguel P, Ellinor PT, Vasan RS, Bruneval P, Markwald RR, Norris RA, Milan DJ, Slaugenhaupt SA, Levine RA, Schott J-J, Hagege AA, MVP-France, Jeunemaitre X, Leducq Transatlantic MITRAL Network. Genetic association analyses highlight biological pathways underlying mitral valve prolapse. Nat Genet. 2015;47:1206–1211.

15. Toomer KA, Yu M, Fulmer D, Guo L, Moore KS, Moore R, Drayton KD, Glover J, Peterson N, Ramos-Ortiz S, Drohan A, Catching BJ, Stairley R, Wessels A, Lipschutz JH, Delling FN, Jeunemaitre X, Dina C, Collins RL, Brand H, Talkowski ME, Del Monte F, Mukherjee R, Awgulewitsch A, Body S, Hardiman G, Hazard ES, da Silveira WA, Wang B, Leyne M, Durst R, Markwald RR, Le Scouarnec S, Hagege A, Le Tourneau T, Kohl P, Rog-Zielinska EA, Ellinor PT, Levine RA, Milan DJ, Schott J-J, Bouatia-Naji N, Slaugenhaupt SA, Norris RA. Primary cilia defects causing mitral valve prolapse. Sci Transl Med. 2019;11.

16. Yu M, Georges A, Tucker NR, Kyryachenko S, Toomer K, Schott J-J, Delling FN, Fernandez-Friera L, Solis J, Ellinor PT, Levine RA, Slaugenhaupt SA, Hagège AA, Dina C, Jeunemaitre X, Milan DJ, Norris RA, Bouatia-Naji N. Genome-Wide Association Study-Driven Gene-Set Analyses, Genetic, and Functional Follow-Up Suggest GLIS1 as a Susceptibility Gene for Mitral Valve Prolapse. Circ Genom Precis Med. 2019;12:e002497.

17. Roselli C, Yu M, Nauffal V, Georges A, Yang Q, Love K, Weng L-C, Delling FN, Maurya SR, Schrölkamp M, Tfelt-Hansen J, Hagège A, Jeunemaitre X, Debette S, Amouyel P, Guan W, Muehlschlegel JD, Body SC, Shah S, Samad Z, Kyryachenko S, Haynes C, Rienstra M, Le Tourneau T, Probst V, Roussel R, Wijdh-Den Hamer IJ, Siland JE, Knowlton KU, Jacques Schott J, Levine RA, Benjamin EJ, Vasan RS, Horne BD, Muhlestein JB, Benfari G, Enriquez-Sarano M, Natale A, Mohanty S, Trivedi C, Shoemaker MB, Yoneda ZT, Wells QS, Baker MT, Farber-Eger E, Michelena HI, Lundby A, Norris RA, Slaugenhaupt SA, Dina C, Lubitz SA, Bouatia-Naji N, Ellinor PT, Milan DJ. Genome-wide association study reveals novel genetic loci: a new polygenic risk score for mitral valve prolapse. Eur Heart J. 2022;43:1668–1680.

18. Haataja TJK, Bernardi RC, Lecointe S, Capoulade R, Merot J, Pentikäinen U. Non-syndromic Mitral Valve Dysplasia Mutation Changes the Force Resilience and Interaction of Human Filamin A. Structure. 2019;27:102–112.e4.

19. Duval D, Lardeux A, Le Tourneau T, Norris RA, Markwald RR, Sauzeau V, Probst V, Le Marec H, Levine R, Schott JJ, Merot J. Valvular dystrophy associated filamin A mutations reveal a new role of its first repeats in small-GTPase regulation. Biochim Biophys Acta. 2014;1843:234–244.

20. Sauls K, de Vlaming A, Harris BS, Williams K, Wessels A, Levine RA, Slaugenhaupt SA, Goodwin RL, Pavone LM, Merot J, Schott J-J, Le Tourneau T, Dix T, Jesinkey S, Feng Y, Walsh C, Zhou B, Baldwin S, Markwald RR, Norris RA. Developmental basis for filamin-A-associated myxomatous mitral valve disease. Cardiovasc Res. 2012;96:109–119.

21. Duval D, Labbé P, Bureau L, Le Tourneau T, Norris RA, Markwald RR, Levine R, Schott J-J, Mérot J. MVP-Associated Filamin A Mutations Affect FlnA-PTPN12 (PTP-PEST) Interactions. J Cardiovasc Dev Dis. 2015;2:233–247.

22. Delling FN, Noseworthy PA, Adams DH, Basso C, Borger M, Bouatia-Naji N, Elmariah S, Evans F, Gerstenfeld E, Hung J, Le Tourneau T, Lewis J, Miller MA, Norris RA, Padala M, Perazzolo-Marra M, Shah DJ, Weinsaft JW, Enriquez-Sarano M, Levine RA. Research Opportunities in the Treatment of Mitral Valve Prolapse: JACC Expert Panel. J Am Coll Cardiol. 2022;80:2331–2347.

23. Delwarde C, Toquet C, Aumond P, Kayvanjoo AH, Foucal A, Le Vely B, Baudic M, Lauzier B, Blandin S, Véziers J, Paul-Gilloteaux P, Lecointe S, Baron E, Massaiu I, Poggio P, Rémy S, Anegon I, Le Marec H, Monassier L, Schott J-J, Mass E, Barc J, Le Tourneau T, Merot J, Capoulade R. Multimodality imaging and transcriptomics to phenotype mitral valve dystrophy in a unique knock-in Filamin-A rat model. Cardiovasc Res. 2023;119:759–771.

24. Zoghbi WA, Adams D, Bonow RO, Enriquez-Sarano M, Foster E, Grayburn PA, Hahn RT, Han Y, Hung J, Lang RM, Little SH, Shah DJ, Shernan S, Thavendiranathan P, Thomas JD, Weissman NJ. Recommendations for Noninvasive Evaluation of Native Valvular Regurgitation: A Report from the American Society of Echocardiography Developed in Collaboration with the Society for Cardiovascular Magnetic Resonance. J Am Soc Echocardiogr. 2017;30:303–371.

25. Constant Dit Beaufils A-L, Huttin O, Jobbe-Duval A, Senage T, Filippetti L, Piriou N, Cueff C, Venner C, Mandry D, Sellal J-M, Le Scouarnec S, Capoulade R, Marrec M, Thollet A, Beaumont M, Hossu G, Toquet C, Gourraud J-B, Trochu J-N, Warin-Fresse K, Marie P-Y, Schott J-J, Roussel J-C, Serfaty J-M, Selton-Suty C, Le Tourneau T. Replacement Myocardial Fibrosis in Patients With Mitral Valve Prolapse: Relation to Mitral Regurgitation, Ventricular Remodeling, and Arrhythmia. Circulation. 2021;143:1763–1774.

26. Morgenthaler S, Thilly WG. A strategy to discover genes that carry multi-allelic or mono-allelic risk for common diseases: a cohort allelic sums test (CAST). Mutat Res. 2007;615:28–56.

27. Keogh RJ. New technology for investigating trophoblast function. Placenta. 2010;31:347–350.

28. Guilluy C, Dubash AD, García-Mata R. Analysis of RhoA and Rho GEF activity in whole cells and the cell nucleus. Nat Protoc. 2011;6:2050–2060.

29. Delwarde C, Toquet C, Aumond P, Kayvanjoo AH, Foucal A, Le Vely B, Baudic M, Lauzier B, Blandin S, Véziers J, Paul-Gilloteaux P, Lecointe S, Baron E, Massaiu I, Poggio P, Rémy S, Anegon I, Le Marec H, Monassier L, Schott JJ, Mass E, Barc J, Le Tourneau T, Merot J, Capoulade R. Multimodality imaging and transcriptomics to phenotype mitral valve dystrophy in a unique knock-in Filamin-A rat model. Cardiovasc Res. 2022;cvac136.

30. Ehrlicher AJ, Nakamura F, Hartwig JH, Weitz DA, Stossel TP. Mechanical strain in actin networks regulates FilGAP and integrin binding to filamin A. Nature. 2011;478:260–263.

31. Nakamura F. FilGAP and its close relatives: a mediator of Rho-Rac antagonism that regulates cell morphology and migration. Biochem J. 2013;453:17–25.

32. Adams DH, Rosenhek R, Falk V. Degenerative mitral valve regurgitation: best practice revolution. Eur Heart J. 2010;31:1958–1966.

33. Cooper MJ, Abinader EG. Family history in assessing the risk for progression of mitral valve prolapse. Report of a kindred. Am J Dis Child. 1981;135:647–649.

34. Trochu JN, Kyndt F, Schott JJ, Gueffet JP, Probst V, Bénichou B, Le Marec H. Clinical characteristics of a familial inherited myxomatous valvular dystrophy mapped to Xq28. J Am Coll Cardiol. 2000;35:1890–1897.

35. Gould RA, Yalcin HC, MacKay JL, Sauls K, Norris R, Kumar S, Butcher JT. Cyclic Mechanical Loading Is Essential for Rac1-Mediated Elongation and Remodeling of the Embryonic Mitral Valve. Current Biology [Internet]. 2016 [cited 2022 Nov 10];26:27–37. Available from: https://linkinghub.elsevier.com/retrieve/pii/S096098221501427X

36. Hale ZE, Sadoshima J. Primary Cilia and Their Role in Acquired Heart Disease. Cells. 2022;11:960.

37. Akilesh S, Suleiman H, Yu H, Stander MC, Lavin P, Gbadegesin R, Antignac C, Pollak M, Kopp JB, Winn MP, Shaw AS. Arhgap24 inactivates Rac1 in mouse podocytes, and a mutant form is associated with familial focal segmental glomerulosclerosis. J Clin Invest. 2011;121:4127–4137.

38. Sanematsu F, Hirashima M, Laurin M, Takii R, Nishikimi A, Kitajima K, Ding G, Noda M, Murata Y, Tanaka Y, Masuko S, Suda T, Meno C, Côté J-F, Nagasawa T, Fukui Y. DOCK180 is a Rac activator that regulates cardiovascular development by acting downstream of CXCR4. Circ Res. 2010;107:1102–1105.

39. Shifrin Y, Arora PD, Ohta Y, Calderwood DA, McCulloch CA. The role of FilGAP-filamin A interactions in mechanoprotection. Mol Biol Cell. 2009;20:1269–1279.

40. Ohta Y, Hartwig JH, Stossel TP. FilGAP, a Rho- and ROCK-regulated GAP for Rac binds filamin A to control actin remodelling. Nat Cell Biol. 2006;8:803–814.

41. Kawaguchi K, Saito K, Asami H, Ohta Y. ADP ribosylation factor 6 (Arf6) acts through FilGAP protein to down-regulate Rac protein and regulates plasma membrane blebbing. J Biol Chem. 2014;289:9675–9682.

42. Misra S, Ghatak S, Moreno-Rodriguez RA, Norris RA, Hascall VC, Markwald RR. Periostin/Filamin-A: A Candidate Central Regulatory Axis for Valve Fibrogenesis and Matrix Compaction. Front Cell Dev Biol. 2021;9:649862.

43. Disse S, Abergel E, Berrebi A, Houot AM, Le Heuzey JY, Diebold B, Guize L, Carpentier A, Corvol P, Jeunemaitre X. Mapping of a first locus for autosomal dominant myxomatous mitral-valve prolapse to chromosome 16p11.2-p12.1. Am J Hum Genet. 1999;65:1242–1251.

44. Freed LA, Acierno JS, Dai D, Leyne M, Marshall JE, Nesta F, Levine RA, Slaugenhaupt SA. A locus for autosomal dominant mitral valve prolapse on chromosome 11p15.4. Am J Hum Genet. 2003;72:1551–1559.

