## Supplementary material for "Isolated prolapse of the posterior mitral valve leaflet: phenotypic refinement, heritability and genetic etiology": Supp. Table 1

**Supplemental Table 1**

| **Proband** | **Gender** | ***ARHGAP24* variants** | ***IFT140* variants** | ***DCHS1* variants** |
|---|---|---|---|---|
| PostMVP1 | Male | p.Thr481Met | - | - |
| PostMVP2 | Female | p.Pro417His | p.Met1164Val | - |
| PostMVP3 | Male | - | p.Arg1433Cys | p.Glu1066Gly |
| PostMVP4 | Male | p.Thr481Met | p.Leu1117Pro | - |
| PostMVP5 | Male | p.Asp92Asn | p.Arg253Gln | - |
| PostMVP6 | Male | - | - | - |
| PostMVP7 | Male | p.Gln671* | - | - |
| PostMVP8 | Female | - | - | - |
| PostMVP9 | Male | - | - | - |
| PostMVP10 | Male | - | - | - |
| PostMVP11 | Male | - | - | - |
| PostMVP12 | Male | - | - | - |
| PostMVP13 | Male | - | - | - |
| PostMVP14 | Male | - | - | p.Ala516Thr |
| PostMVP15 | Female | - | - | - |
| PostMVP16 | Male | - | p.Asp443Asn | - |
| PostMVP17 | Male | - | p.Ala473Thr | - |
| PostMVP18 | Female | - | - | - |
| PostMVP19 | Male | - | p.Arg280Gln | - |
| PostMVP20 | Male | - | p.Arg253Gln | p.Arg917Gln |

*ARHGAP24* : ENST00000395184; IFT140: ENST00000426508; DCHS1: ENST00000299441
