## Supplementary material for "Isolated prolapse of the posterior mitral valve leaflet: phenotypic refinement, heritability and genetic etiology": Supp. Table 2

**Supplemental Table 2**

| GENE | GENE_ID | p-value (CAS | Cases | Controls | OR |
|---|---|---|---|---|---|
| ***IFT140*** | **ENSG00000[illegible]** | **9.55E-07** | **8** | **33** | **16.4624** |
| ***ARHGAP24*** | **ENSG00000[illegible]** | **0.00030555** | **5** | **24** | **11.4584** |
| ***ZNF221*** | **ENSG00000[illegible]** | **0.00061702** | **4** | **15** | **13.8709** |
| *SEMA4G* | ENSG000000 | 0.00157039 | 4 | 20 | 10.3683 |
| *TELO2* | ENSG000001 | 0.00184191 | 4 | 21 | 9.866 |
| *SALL4* | ENSG000001 | 0.00184191 | 4 | 21 | 9.866 |
| *LPHN2* | ENSG000001 | 0.00325835 | 4 | 25 | 8.2588 |
| *FANCM* | ENSG000001 | 0.00656866 | 4 | 31 | 6.6206 |
| *LTBP1* | ENSG000000 | 0.0080419 | 4 | 33 | 6.2053 |
| *NLRX1* | ENSG000001 | 0.00972067 | 4 | 35 | 5.8388 |
| *ERCC6* | ENSG000002 | 0.011698 | 5 | 59 | 4.489 |
| *CASZ1* | ENSG000001 | 0.01264871 | 4 | 38 | 5.3604 |
| *MYLK* | ENSG000000 | 0.01264871 | 4 | 38 | 5.3604 |
| *DNAH11* | ENSG000001 | 0.0203777 | 6 | 95 | 3.426 |
| *CSPG4* | ENSG000001 | 0.02986788 | 4 | 50 | 4.0189 |
| *STAB1* | ENSG000000 | 0.03366509 | 4 | 52 | 3.8554 |
| *NEB* | ENSG000001 | 0.03399852 | 6 | 107 | 2.9946 |
| *MYO15A* | ENSG000000 | 0.05163876 | 5 | 88 | 2.9039 |
| *PLEC* | ENSG000001 | 0.0635198 | 7 | 144 | 2.6585 |
| *UBR4* | ENSG000001 | 0.06521622 | 4 | 65 | 3.0363 |
| *IGFN1* | ENSG000001 | 0.07113332 | 4 | 67 | 2.9387 |
| *FAT1* | ENSG000000 | 0.0722518 | 5 | 97 | 2.6043 |
| *DNAH12* | ENSG000001 | 0.07419695 | 4 | 68 | 2.8916 |
| *CACNA1H* | ENSG000001 | 0.07419695 | 4 | 68 | 2.8916 |
| *RP11-1055B8* | ENSG000001 | 0.10484329 | 4 | 77 | 2.5254 |
| *HYDIN* | ENSG000001 | 0.13230716 | 4 | 84 | 2.2947 |
| *SRRM2* | ENSG000001 | 0.27450103 | 4 | 97 | 1.9543 |
| *GPR98* | ENSG000001 | 0.29842469 | 4 | 105 | 1.7866 |
| *SYNE2* | ENSG000000 | 0.32747715 | 4 | 113 | 1.6427 |
| *DNAH17* | ENSG000001 | 0.33945924 | 4 | 116 | 1.5938 |
| *LAMA5* | ENSG000001 | 0.52543866 | 4 | 128 | 1.4212 |
| *TTN* | ENSG000001 | 0.65171667 | 11 | 406 | 1.3542 |
| *MUC16* | ENSG000001 | 0.7985722 | 6 | 228 | 1.1802 |
