## Supplementary material for "Isolated prolapse of the posterior mitral valve leaflet: phenotypic refinement, heritability and genetic etiology": Supp. Table 3

### Supplemental Table 3

| Chr | Pos (GRCh37) | Ref | Alt | Carriers | dbSNP | AAChange.ensGene | SIFT_pred | Polyphen2_HDIV_pred | LRT_pred | CADD_phred | GERP++_RS | Interpro_domain | gnomAD_genome_NFE |
|---|---|---|---|---|---|---|---|---|---|---|---|---|---|
| chr4 | 86491702 | A | G | 1 BiMVP patient, 2 controls | rs150698221 | ENSG00000138639:**ENST00000395184:exon2:c.A8G:p.E3G**,ENSG00000138639:ENST00000503995:exon2:c.A8G:p.E3G | D | P | D | 24.6 | 5.81 | . | 0.0005 |
| chr4 | 86491861 | A | G | 1 control | rs149415202 | ENSG00000138639:**ENST00000395184:exon2:c.A167G:p.E56G**,ENSG00000138639:ENST00000503995:exon2:c.A167G:p.E56G | D | D | D | 25.4 | 5.91 | PH domain-like\|Pleckstrin homology domain | 6.668e-05 |
| chr4 | 86643057 | G | A | 1 BiMVP patient | rs144785317 | ENSG00000138639:**ENST00000395184:exon3:c.G200A:p.G67E**,ENSG00000138639:ENST00000503995:exon3:c.G200A:p.G67E | D | P | D | 27.7 | 4.97 | PH domain-like\|Pleckstrin homology domain | 0.0010 |
| chr4 | 86643074 | C | T | 1 BiMVP patient | rs748396958 | ENSG00000138639:**ENST00000395184:exon3:c.C217T:p.H73Y**,ENSG00000138639:ENST00000503995:exon3:c.C217T:p.H73Y | D | D | D | 23.8 | 4.97 | PH domain-like\|Pleckstrin homology domain | . |
| chr4 | 86844806 | G | A | **PostMVP5**, 1 BiMVP patient, 3 controls | rs571519833 | ENSG00000138639:ENST00000514229:exon2:c.G19A:p.D7N,ENSG00000138639:**ENST00000395184:exon4:c.G274A:p.D92N**,ENSG00000138639:ENST00000503995:exon4:c.G274A:p.D92N | D | D | D | 25.4 | 06.03 | PH domain-like\|Pleckstrin homology domain;Pleckstrin homology domain | . |
| chr4 | 86844816 | G | A | 1 PostMVP patient, 1 control | rs145434155 | ENSG00000138639:ENST00000514229:exon2:c.G29A:p.R10Q,ENSG00000138639:**ENST00000395184:exon4:c.G284A:p.R95Q**,ENSG00000138639:ENST00000503995:exon4:c.G284A:p.R95Q | T | B | D | 24.2 | 05.02 | PH domain-like\|Pleckstrin homology domain;Pleckstrin homology domain | 0.0006 |
| chr4 | 86844840 | A | G | 1 control | . | ENSG00000138639:ENST00000395183:exon2:c.A23G:p.Y8C,ENSG00000138639:ENST00000514229:exon2:c.A53G:p.Y18C,ENSG00000138639:**ENST00000395184:exon4:c.A308G:p.Y103C**,ENSG00000138639:ENST00000503995:exon4:c.A308G:p.Y103C,ENSG00000138639:ENST00000512201:exon4:c.A23G:p.Y8C | T | D | D | 28.9 | 06.03 | PH domain-like\|Pleckstrin homology domain | . |

| | | | | | | | | | | | | | |
|---|---|---|---|---|---|---|---|---|---|---|---|---|---|
| chr4 | 86844880 | G | T | 1 control | rs766244542 | ENSG00000138639:ENST00000395183:exon2:c.G63T:p.W21C,ENSG00000138639:ENST00000514229:exon2:c.G93T:p.W31C,ENSG00000138639:**ENST00000395184:exon4:c.G348T:p.W116C**,ENSG00000138639:ENST00000503995:exon4:c.G348T:p.W116C,ENSG00000138639:ENST00000512201:exon4:c.G63T:p.W21C | D | D | D | 33 | 6.17 | PH domain-like\|Pleckstrin homology domain | 0.0002 |
| chr4 | 86852171 | A | G | 1 control | . | ENSG00000138639:ENST00000264343:exon1:c.A88G:p.K30E | D | B | . | 20.7 | 05.09 | PH domain-like\|Pleckstrin homology domain | . |
| chr4 | 86915976 | G | C | 1 control | . | ENSG00000138639:ENST00000264343:exon6:c.G890C:p.G297A,ENSG00000138639:ENST00000395183:exon7:c.G884C:p.G295A,ENSG00000138639:ENST00000514229:exon7:c.G914C:p.G305A,ENSG00000138639:**ENST00000395184:exon9:c.G1169C:p.G390A** | T | B | D | 10.49 | 4.57 | Rho GTPase activation protein\|Rho GTPase-activating protein domain | . |
| chr4 | 86916057 | C | A | **PostMVP2** | rs756950646 | ENSG00000138639:ENST00000264343:exon6:c.C971A:p.P324H,ENSG00000138639:ENST00000395183:exon7:c.C965A:p.P322H,ENSG00000138639:ENST00000514229:exon7:c.C995A:p.P332H,ENSG00000138639:**ENST00000395184:exon9:c.C1250A:p.P417H** | D | D | D | 28.1 | 5.6 | Rho GTPase activation protein\|Rho GTPase-activating protein domain | . |
| chr4 | 86916193 | C | A | 1 control | rs142672228 | ENSG00000138639:ENST00000264343:exon6:c.C1107A:p.S369R,ENSG00000138639:ENST00000395183:exon7:c.C1101A:p.S367R,ENSG00000138639:ENST00000514229:exon7:c.C1131A:p.S377R,ENSG00000138639:**ENST00000395184:exon9:c.C1386A:p.S462R** | T | B | N | 17.09 | 3.28 | . | 6.663e-05 |
| chr4 | 86916212 | G | A | 1 control | . | ENSG00000138639:ENST00000264343:exon6:c.G1126A:p.G376S,ENSG00000138639:ENST00000395183:exon7:c.G1120A:p.G374S,ENSG00000138639:ENST00000514229:exon7:c.G1150A:p.G384S,ENSG00000138639:**ENST00000395184:exon9:c.G1405A:p.G469S** | D | B | D | 22.9 | 5.72 | . | . |
| chr4 | 86916232 | C | G | 1 control | rs141031654 | ENSG00000138639:ENST00000264343:exon6:c.C1146G:p.H382Q,ENSG00000138639:ENST00000395183:exon7:c.C1140G:p.H380Q,ENSG00000138639:ENST00000514229:exon7:c.C1170G:p.H390Q,ENSG00000138639:**ENST00000395184:exon9:c.C1425G:p.H475Q** | T | B | D | 10.41 | 3.96 | . | 0.0001 |

| | | | | | | | | | | | | | |
|---|---|---|---|---|---|---|---|---|---|---|---|---|---|
| chr4 | 86916249 | C | T | **PostMVP1, PostMVP4**, 1 PostMVP patient, 1 BiMVP patient, 8 controls | rs147870358 | ENSG00000138639:ENST00000264343:exon6:c.C1163T:p.T388M,ENSG00000138639:ENST00000395183:exon7:c.C1157T:p.T386M,ENSG00000138639:ENST00000514229:exon7:c.C1187T:p.T396M,ENSG00000138639:**ENST00000395184:exon9:c.C1442T:p.T481M** | D | P | N | 23.6 | 4.86 | . | 0.0011 |
| chr4 | 86916254 | C | T | 1 control | rs777874622 | ENSG00000138639:ENST00000264343:exon6:c.C1168T:p.R390C,ENSG00000138639:ENST00000395183:exon7:c.C1162T:p.R388C,ENSG00000138639:ENST00000514229:exon7:c.C1192T:p.R398C,ENSG00000138639:**ENST00000395184:exon9:c.C1447T:p.R483C** | D | D | D | 27.1 | 4.87 | . | 6.667e-05 |
| chr4 | 86916303 | G | A | 1 control | rs115120658 | ENSG00000138639:ENST00000264343:exon6:c.G1217A:p.R406Q,ENSG00000138639:ENST00000395183:exon7:c.G1211A:p.R404Q,ENSG00000138639:ENST00000514229:exon7:c.G1241A:p.R414Q,ENSG00000138639:**ENST00000395184:exon9:c.G1496A:p.R499Q** | D | D | D | 23.2 | 05.02 | . | 0.0001 |
| chr4 | 86921639 | C | T | **PostMVP7** | rs777805344 | ENSG00000138639:ENST00000264343:exon7:c.C1732T:p.Q578X,ENSG00000138639:ENST00000395183:exon8:c.C1726T:p.Q576X,ENSG00000138639:ENST00000514229:exon8:c.C1756T:p.Q586X,ENSG00000138639:**ENST00000395184:exon10:c.C2011T:p.Q671X** | . | . | D | 42 | 5.56 | . | . |
| chr4 | 86921668 | G | C | 1 PostMVP patient | rs200818558 | ENSG00000138639:ENST00000264343:exon7:c.G1761C:p.M587I,ENSG00000138639:ENST00000395183:exon8:c.G1755C:p.M585I,ENSG00000138639:ENST00000514229:exon8:c.G1785C:p.M595I,ENSG00000138639:**ENST00000395184:exon10:c.G2040C:p.M680I** | T | B | D | 21.2 | 5.56 | . | 0.0004 |
