## Supplemental for "Isolated prolapse of the posterior mitral valve leaflet: phenotypic refinement, heritability and genetic etiology"

**SUPPLEMENTARY APPENDIX**

### SUPPLEMENTAL METHODS

#### Echocardiography

All values were obtained from the mean of 3 beats and from the mean of 5 to 10 beats in patients with atrial fibrillation. The mitral valve (MV) apparatus was comprehensively assessed. In the parasternal long axis, anterior and posterior leaflets lengths and thickness tips, and mitral annulus diameter as well, were measured in diastole. The maximal distance between each leaflet and the annulus plane at end-systole was measured and expressed positively (normal closing) or negatively (prolapse). Chordal lengths were measured from the papillary muscle (PM) tip to leaflet tips in systole by tilting the probe toward the posterior commissure (posterior *chordae tendienae*) or toward the anterior commissure (anterior *chordae tendieneae*). In a modified apical left ventricular (LV) 2-chambers view showing both PM tips the distance of the PM tips to mitral annulus line (PM tip position) was measured at end-diastole and expressed as the ratio of the PM tip-mitral annulus distance to LV long axis length.

Quantification of regurgitation was based at least on 2 different methods (proximal isovelocity surface area, continuity equation, LV volumes) and results were averaged. Mitral regurgitation was classified into trace/mild, or moderate/severe according to a regurgitant volume < 45 mL/beat (trace/mild) or ≥ 45 mL/beat (moderate/severe).

For operated relatives valve lesion characteristics and degree of regurgitation were obtained from pre and per-operative assessment.

The diagnosis of mitral valve prolapse (MVP) was based on a superior displacement > 2 mm above the mitral annulus line of at least one leaflet in the PLA view^1,2^. Prodromal forms were defined as the conjunction of a minimal systolic displacement of the posterior MV leaflets between 0.1 to 2 mm and an AAC (abnormal anterior coaptation). Patients with either MVP or MVP prodromal were considered affected by the disease as in prior genetic studies.

#### Whole-genome sequencing

##### Sequencing and alignment

Whole genome sequencing was performed by the “Centre National de Recherche en Génomique Humaine” (CNRGH, Institut de Biologie François Jacob, CEA, Evry, France). After a complete quality control, 1µg of genomic DNA was used to prepare a library for whole genome sequencing, using the Illumina TruSeq DNA PCR-Free Library Preparation Kit, according to the manufacturer's instructions. After normalization and quality control, qualified libraries were sequenced on a NovaSeq6000 platform from Illumina (Illumina Inc., CA, USA), as paired-end 150 bp reads. Libraries were pooled in order to reach an average sequencing depth of 30x for each sample. Sequence quality parameters have been assessed throughout the sequencing run and standard bioinformatics analysis of sequencing data was based on the Illumina pipeline to generate a FASTQ file for each sample.

Raw sequence reads were aligned to the human reference genome (GRCh37) using BWA-MEM (version 0.7.15)^3^. GATK 4.2.0.0 was used for indel realignment and base recalibration, following GATK DNA Best Practices. The mean sequencing depth ranged from 34× to 57×.

##### Calling, filtering and burden analysis

Variants were called with HaplotypeCaller in GVCF mode.

The following filters were applied and excluded variants:

- with more than 2 alternative alleles using bcftools^4^,
- with an alternative allele and a genotype quality (GQ) lower than 50 using jvarkit/vcffilterjdk^5^,
- singletons with an allele balance below 0.2 or above 0.8 jvarkit/vcffilterjdk

Variants were considered as ‘rare’ if the frequency was <0.2% in the Non-Finnish European population in gnomAD v2.1 and as ‘protein-altering’ if they were annotated with at least one of the following SO terms: “transcript_ablation” (SO:0001893), “splice_donor_variant” (SO:0001575), “splice_acceptor_variant” (SO:0001574), “stop_gained” (SO:0001587), “frameshift_variant” (SO:0001589), “stop_lost” (SO:0001578), “start_lost” (SO:0002012), “inframe_insertion” (SO:0001821), “inframe_deletion” (SO:0001822), “missense_variant” (SO:0001583), “transcript_amplification” (SO:0001889), or “protein_altering_variant” (SO :0001818). Usually, variants are considered as rare when they present a frequency lower than 1% in the general population. Knowing the high prevalence of MVP (~2%)we set the frequency cut-off of 0.2% for variants of interest regarding the pathology. The burden analysis was performed using the cohort allelic sums test (CAST)^6^. We focused on genes with at least 4 variants of interest in MVP patients. Visual validation of genetic variants was performed with the integrative genome viewer^7^ using Bam files.

#### Sanger Sequencing

 All relevant variants identified in individuals sequenced were manually reviewed by visual inspection of sequence reads using the Integrative Genomics Viewer ^7^. When mentioned in the manuscript, Sanger sequencing was performed to validate *in-silico* data. After amplification by PCR, excess primers were removed from the amplified fragments using exoSAP (Amersham Biosciences, Piscataway, NJ) and sequenced with a dye-terminator cycle-sequencing system (ABI PRISM 3730, Perkin-Elmer Applied Biosystems, Foster City, Calif).

#### Cellular model

Hek293T cells were cultured in DMEM supplemented with 10% fetal calf serum and L-glutamine.

#### Plasmids

The pCMV5 N-terminally HA-tagged FilGAP (HA-FilGAP-WT) was a gift of Dr Yasutaka Ohta (Division of Cell Biology, Kitasato University, Japan). Mutations were generated by directed mutagenesis (QuikChange II XL Site-Directed Mutagenesis Kit – Agilent) following the manufacturer’s specifications. All the constructs were verified by Sanger sequencing (data not shown). Cells were transfected with FilGAP constructs plasmids using Lipofectamine (Invitrogen) or Genecellin (BiocellChallenge) according to the manufacturers’ specifications.

#### RT-PCR

mRNA was reverse transcribed and amplified using the High-Capacity cDNA Reverse transcription kits (Applied Biosystems) and amplified by PCR following the manufacturers recommendations. The following forward (F) and reverse (R) isoforms specific *ARHGAP24* primers were used:

iso1-F:5’-CAATGACTCCACGGAGAACC-3’

iso1-R:5’-TCCCTGGGTTCTCTTCATTG-3’

iso2-F: 5’-AAACCGGGTTCAGAACTTCA-3’

iso2-R: 5’-CCCACAGTCAAAGGCATCTT-3’

iso3-F: 5’-TGGGATGGGAGGATACTGAC-3’

iso3-R: 5’-ATATGACTCGGCGGATTGAC-3’

iso4-F: 5’-CTGAAGTGTATGTTTGTGCAAG-3’

For *GAPDH*

GAPDH-F: 5’-TTCATTGACCTCAACTACATGGT-3’

GAPDH-R: 5’-CTCAGTGTAGCCCAGGATGCCCTT-3’.

#### Co-immunoprecipitation and immunoblotting

Cells transfected with FilGAP-HA and other tagged FilGAP (EGFP) were lysed in NETF buffer containing: 100 mM NaCl, 2 mM EDTA, 50 mM Tris pH 7.5, 50 mM NaF, 1 % NP- 40, 1 mM PMSF, 1 mM Na3VO4, 1X protease inhibitor cocktail (Roche) and lysates clarified by centrifugation (15,000   g for 15 min at 4 C). The cell lysates (500 μg) were incubated with 6 μg of anti-HA for 2h at 4°C and then with 30 μl of protein A conjugated beads (Dynabeads, Invitrogen) for 1h at 4 C. The immunoprecipitates were washed four times with NETF buffer and separated by SDS-PAGE transferred to nitrocellulose membrane (Bio-Rad Transblot). Immunoblots probed with appropriate antibodies and revealed using enhanced chemiluminescence (ECL) kit (GE Healthcare). Chemiluminescence signals were quantified using an Imager system (Roche Diagnostic) and the data normalized with respect to GAPDH.

#### siRNA

 To deplete endogenous FilGAP, siRNA oligonucleotide duplexes targeting the sequence 5′-AAGATAGAGTATGAGTCCAGGATAA-3′ (nt 1975–1999 of FilGAP) were used (3). Control siRNA duplexes targeting GFP were used (sense 5’-GCAAGCUGACCCUGAAGUUCAU-3’, antisense 5’-GAACUUCAGGGU CAGCUUGCCG-3’). The cells were transfected according to the suppliers’ guidelines (Eurogentec) and used 48 hours post-transfection.

#### Cellular adhesion and spreading monitoring

The impedance measurement technology of the xCELLigence system was used to monitor cell adhesion and spreading as previously described ^8,9^. 10 000 cells per well were plated into 96-wells E-Plates (Roche Diagnostics, GmbH), placed on the Real Time Cell Analyzer and incubated at 37°C in a 5% CO_2_ incubator. Cell adhesion and spreading were measured and expressed as a C*ell Index* (CI) according to the manufacturer's guidelines. Impedance measurements were taken every 1 min for 3 hours. The slope of CI changes (dCI/dt) were calculated between t_30min_ and t_1h30min_.

#### Glutathione-S-transferase (GST) protein purification and GST pull-down assays.

GST-PAK1 containing the Cdc42/Rac1 Interactive Binding (Crib) region of p21 activated kinase were produced in Bl21 E.Coli treated overnight with 1 mM IPTG at 25°C. The GST-fusion proteins were purified using Glutathione Agarose 4B beads (Macherey-Nagel). Transfected Hek293 cells grown for 2-4 hrs after seeding and lysed. Cell lysates were centrifuged at 15,000 ×g for 15 min at 4°C. 500 µg of cleared cell-lysates were incubated with GST-tagged proteins (30µg) and rotated (18 rpm) for 1h at 4°C. The beads were washed four times with cell lysis buffer and bound proteins separated by SDS-PAGE. Bound Rac1 was detected by immunoblotting as described above.

### SUPPLEMENTAL TABLES

### **Supplemental Table 1: Rare protein-altering variants in MVP-related genes in 20 PostMVP probands.**

Variants in *ARHGAP24* correspond to the transcript ENST00000395184, variants in *DCHS1* to ENST00000299441.5 and variants in *IFT140*: ENST00000426508.7.

#### Supplemental Table 2: Enrichment in rare variants in 20 PostMVP probands (n = 20) vs. controls (n= 856)

#### Supplemental Table 3: Rare variants in *ARHGAP24* in 284 MVP patients and 856 controls

#### Annotations were obtained from wANNOVAR (https://wannovar.wglab.org) in October 2022.

### SUPPLEMENTAL FIGURES

#### Supplemental figure 1: Flow chart patients’ selection

#### Supplemental figure 2: Pedigree trees of 20 PostMVP familial cases

Pedigree tree of PostMVP-familial cases. PostMVP patients are indicated by filled black symbols, patients with prodromal forms of MVP with gray symbols, BiMVP patients with hashed symbols and white symbols depict individuals with normal mitral valve. The “?” sign indicates patients not examined by the clinicians. Arrows show index cases. ‘*’ corresponds to patients with mitral valve surgery.

#### Supplemental figure 3: Co-segregation analysis of *DCHS1* rare variants in PostMVP familial cases

Pedigree tree of PostMVP-familial cases carrying *DCHS1* variants. PostMVP patients are indicated by filled black symbols, patients with prodromal forms of MVP with gray symbols, BiMVP patients with hashed symbols and white symbols depict individuals with normal mitral valve. The “?” sign indicates patients not examined by the clinicians. *DCHS1* variant carriers and non-carriers are indicated in blue below symbols (Wt = wild type). Arrows show index cases. ‘*’ corresponds to patients with mitral valve surgery.

#### Supplemental figure 4: *ARHGAP24* variants in PostMVP and BiMVP patients

Variants are presented using the canonical FilGAP protein isoform (ENST00000395184 -ENSP00000378611, 748 amino acids). Variants in red were identified in PostMVP patients only, blue ones were found in BiMVP patients only and variants in green were found in both groups. Variants detected in PostMVP probands (familial cases) are indicated in bold. The p.Thr481Met variant was detected in 2 PostMVP probands, 1 PostMVP sporadic patient, and 1 BiMVP patient.

#### Supplemental figure 5: ARHGAP24 knock down in zebrafish

A) Sequences of the two morpholinos used in the study and of the primers used to quantify

extinction by QPCR.

B) Efficacy of the ARHGAP24-ortholog knock down

C) The survival of the fishes was not significantly modified morpholinos up to 140 Hrs post fertilization.

#### Supplemental figure 6: *ARHGAP24* expression in tissues

**A)** *ARHGAP24* gene expression in human tissues (source: GTEX <https://gtexportal.org/home/gene/ARHGAP24>)

**B)** Tissue expression profile of FilGAP determined by RT-PCR in brain, kidney and mitral valve mRNA. The 4 isoforms of FilGAP are expressed in Mitral Valve but FilGAP isoform 1 is the most expressed isoform compared to kidney and brain.

**C)** Immuno-histological localization of FilGAP in a control human mitral valve posterior leaflet. FilGAP is detected in both endothelial (arrows) and interstitial cells (arrow heads).

#### Supplemental figure 7: Efficacy of ARHGAP24 silencing with si-RNA

The blot image illustrates the ~80% extinction of endogenous FilGAP obtained using the siRNA directed against ARHGAP24.

### SUPPLEMENTAL REFERENCES

1.	Adams DH, Rosenhek R, Falk V. Degenerative mitral valve regurgitation: best practice revolution. *Eur Heart J* 2010;**31**:1958–1966.

2.	Freed LA, Levy D, Levine RA, Larson MG, Evans JC, Fuller DL, Lehman B, Benjamin EJ. Prevalence and clinical outcome of mitral-valve prolapse. *N Engl J Med* 1999;**341**:1–7.

3.	Li H, Durbin R. Fast and accurate long-read alignment with Burrows-Wheeler transform. *Bioinformatics* 2010;**26**:589–595.

4.	Danecek P, Bonfield JK, Liddle J, Marshall J, Ohan V, Pollard MO, Whitwham A, Keane T, McCarthy SA, Davies RM, Li H. Twelve years of SAMtools and BCFtools. *GigaScience* 2021;**10**:giab008.

5.	Lindenbaum P. JVarkit: java-based utilities for Bioinformatics. 2015:0 Bytes.

6.	Morgenthaler S, Thilly WG. A strategy to discover genes that carry multi-allelic or mono-allelic risk for common diseases: a cohort allelic sums test (CAST). *Mutat Res* 2007;**615**:28–56.

7.	Thorvaldsdóttir H, Robinson JT, Mesirov JP. Integrative Genomics Viewer (IGV): high-performance genomics data visualization and exploration. *Brief Bioinformatics* 2013;**14**:178–192.

8.	Duval D, Lardeux A, Le Tourneau T, Norris RA, Markwald RR, Sauzeau V, Probst V, Le Marec H, Levine R, Schott JJ, Merot J. Valvular dystrophy associated filamin A mutations reveal a new role of its first repeats in small-GTPase regulation. *Biochim Biophys Acta* 2014;**1843**:234–244.

9.	Keogh RJ. New technology for investigating trophoblast function. *Placenta* 2010;**31**:347–350.
